# Unraveling the Genetic Overlap Between Parkinson’s Disease and Schizophrenia Through Genome-wide Association and Cell-Type Specific Transcriptomic Analysis

**DOI:** 10.64898/2026.01.06.26343442

**Authors:** Wenhua Sun, Mohammad Dehestani, Alice Braun, Nasser Karmali, Isabel Wurster, Benjamin Röben, Roswitha Kemmner, Kathrin Brockmann, Manu Sharma, Marina Mitjans, Fabian Streit, Thomas Gasser, Vikas Bansal

## Abstract

**Background:** Parkinson’s disease (PD) and schizophrenia (SZ), while clinically distinct, exhibit overlapping symptoms and neurobiological features. Emerging genetic evidence suggests a shared heritable component between the two disorders. In this study, we aim to identify novel genetic loci common to both PD and SZ and to further investigate the convergent molecular mechanisms that may underlie their shared pathophysiology.

**Methods:** We analyzed large-scale genome-wide association studies (GWAS) on SZ (55,193 cases and 74,132 controls) and PD (56,306 cases and 1,417,791 controls) using a proxy-phenotype method combined with a Bayesian statistical approach to evaluate overlap in common genetic variants and enhance statistical power for genetic discovery. To elucidate the biological mechanisms underlying newly identified shared genetic loci associated with PD and SZ, we leveraged single-cell RNA sequencing (scRNA-seq) data from induced pluripotent stem cell (iPSC)-derived dopaminergic neurons generated by the FOUNDIN-PD consortium (416,216 high-quality single-cells from PD patients). We performed differential gene expression analysis, and gene ontology (GO) enrichment analysis to systematically investigate the functional relevance of one shared loci in a disease-relevant cellular context. The polygenic risk score (PRS) model was constructed by stratifying overlapping SNPs based on effect directions to predict PD clinical outcomes in the Tuebingen Parkinson cohort (TUEPAC). MAGMA gene-set enrichment analysis was used to identify the cell types enriched for each SNP set.

**Results:** We identified five SNPs with concordant effect directions and nine SNPs with discordant effect directions shared between PD and SZ. Of these fourteen SNPs, eleven were in linkage disequilibrium with previously reported PD GWAS hits. In addition, we identified three novel shared SNPs (rs2240921, rs11649804, and rs9607782) associated with both PD and SZ. Notably, a missense variant in *RAI1* (rs11649804) emerged as a shared genetic signal between the two disorders. Differential expression analysis and gene ontology (GO) enrichment analysis implicated *RAI1* in mitochondrial dysfunction-related biological processes, highlighting its potential role in PD pathogenesis. In addition, *PD_discordant* PRS was associated with non-motor symptoms and uniquely associated with genes expressed in brain vascular cell types.

**Conclusion:** In summary, our study provides novel insights into the shared genetic architecture of PD and SZ, highlighting previously unrecognized loci with potential cross-disorder relevance. Functional characterization of *RAI1* loci suggests that mitochondrial dysfunction may represent convergent biological mechanisms underlying both neurodegenerative and psychiatric pathologies. Moreover, variants with opposing cross-disorders effects may capture biological pathways specifically contributing to non-motor heterogeneity. These findings advance our understanding of the molecular interplay between PD and SZ and open new avenues for the identification of shared therapeutic targets.

## Introduction

Parkinson’s disease (PD) is the second most common neurodegenerative disorder, affecting around 1%-2% of the population over 65 [1]; it is characterized by the loss of dopaminergic neurons in substantia nigra pars compacta and pathological accumulation of a-synuclein(a-syn) protein in remaining neurons [2]. Schizophrenia (SZ) is a serious psychiatric illness that commonly strikes people in adolescence or early adulthood, and it is related to an overactive dopaminergic system [3], therefore both the disorders have been linked to dopaminergic system dysfunction. At the transcriptional level, previous MAGMA-based cell-type enrichment analyses have linked PD and SZ genetic risk to genes exhibiting dopaminergic neuron-specific expression patterns, based on the human substantia nigra (SN) transcriptomic cellular atlas [4]. The two diseases have a degree of overlapping symptomatology. Clinically, motor dysfunctions in PD include rigidity, rest tremor, bradykinesia, postural instability, and a multitude of non-motor features (cognitive impairment, depression, symptoms of psychosis, such as illusions, hallucinations, and delusions, etc) are often accompanying PD [2, 5]. SZ symptoms are commonly divided into three domains: positive symptoms (hallucinations, delusions), negative symptoms (reduced speech output, motivation), and cognitive impairment [3]. Besides, parkinsonism was reported to exist in unmedicated schizophrenia patients [6]. Additionally, late-life PD risk has been reported to be increased in patients with SCZ [7]. The increased PD risk may arise from two factors: the potential risk-modifying effects of dopamine-blocking medications or heightened vulnerability of the dopamine system due to fluctuating dopamine imbalances during different stages [7, 8].

In addition to dopaminergic dysfunction, non-dopaminergic mechanisms also contribute to non-motor symptoms of PD [5, 9, 10] and SZ [11]. In SZ, serotonin (5-hydroxytryptamine) 2A (5-HT2A) receptors can form a functional complex with metabotropic glutamate receptor 2 (mGlu2) in the frontal cortex [11], and 5-HT2A hyperactivity has been linked to positive symptoms such as hallucinations and delusions [12]. Likewise, increasing evidence supports serotonergic dysfunction in PD [13–17], particularly in relation to non-motor symptoms including depression, fatigue, weight and appetite changes, and visual hallucinations [18, 19].

The latest SZ genome-wide association study (GWAS) has reported a SNP-based heritability of 0.24, which is attributable to common genetic variants and identified 313 independent SNPs [20]. A recent PD GWAS identified 90 independent genome-wide significant loci with an SNP-based heritability estimated at 0.22 [21]. These findings collectively underscore the polygenic architecture of both disorders, wherein many common genetic variants with small effect size cumulatively contribute to disease susceptibility. In addition, a previous study identified SZ GWAS risk genes that were dysregulated in developing SZ glutamatergic neurons, further supporting the involvement of glutamatergic pathways in SZ [22].

Several lines of evidence support a shared genetic background between PD and SZ. Earlier research found that a per standard deviation increase in the allelic burden derived from SZ GWAS risk variants was significantly associated with PD status (odds ratio = 1.05) [23]. In a more recent two-sample Mendelian randomization analysis using large-scale GWAS summary statistics, genetic variants increasing the risk of PD were found to contribute to increased susceptibility to SZ [24]. Notably, another study using non-overlapping GWAS summary statistics for PD and SZ to identify shared common risk variants through a conditional/conjunctional false discovery rate statistical framework [25]. Besides, patients with 22q11.2 deletion syndrome have an increased risk of both SZ (30% prevalence) and early-onset PD [26, 27].

In the current study, we aim to identify shared novel common genetic variants between PD and SZ by applying a proxy-phenotype method (PPM) [28, 29] combined with Bayesian statistical framework, using the large non-overlapping GWAS summary statistics. Furthermore, we explored the underlying mechanisms of these shared novel genetic variants at the transcriptional level in PD through single-cell RNA sequencing data from FOUNDIN-PD [30] and human post-mortem brain single-nuclei RNA sequencing dataset [31]. Although previous studies have laid critical groundwork by identifying shared genetic variants between PD and SZ, further investigation using larger and more recent GWAS may help refine shared genetic variants. Moreover, integrating GWAS findings with single-cell transcriptomic data may provide biological context for the implicated loci. Therefore, in this study, we aimed to identify additional shared genetic variants between PD and SZ and to explore their potential functional relevance using single-cell RNA sequencing data.

## Methods

### GWAS data

We obtained GWAS results from individuals of European ancestry in the form of summary statistics (coordinates, minor allele frequency, effect size, standard error and p value) on PD [21] and SZ [20]. PD GWAS, integrating the dataset from 23andMe, includes 56,306 cases and 1,417,791 controls with 7,200,892 SNPs. SZ GWAS includes 55,193 cases and 74,132 controls with 7,659,767 SNPs. We conducted a systematic review of the source cohorts used in the GWAS for both PD and SZ. We excluded cohorts of WTCCC2 cohort and re-ran the core SZ GWAS meta-analysis aggregated by the Psychiatric Genomics Consortium, using 50,358 cases and 68,967 controls. The custom meta-analysis was carried out with METAL using the inverse variance weighting method[32] as implemented in RICOPILI [33]. This re-analysis served as a sensitivity analysis to ensure that no overlapping case/control participants were included between the PD and SZ studies (Supplementary Document 1). The 2025 PD GWAS was not used in the primary analysis as it was not peer-review published at the time, and the summary statistics was not available at the beginning of analysis. However, we used it to reconfirm the downstream results.

To perform quality control (QC) on the GWAS summary statistics, we utilized MungeSumstats 1.10.1 (a Bioconductor R package) [34] with the following workflow: (1) Data Standardization: The columns names were unified in the outputs files (SNP, the single nucleotide polymorphism reference ID; CHR, chromosome; BP, Base-pair position; A1, non-effect allele; A2, effect allele; FRQ, minor allele frequency (MAF) of the SNP; BETA, effect size estimate relative to the alternative allele; SE, standard error; P, unadjusted P-value; N, sample size). (2) Quality control includes: a. Removal of all non-biallelic SNPs to exclude multi-allelic or ambiguous variants. b. Dropping SNPs with non-standard alleles (i.e. not A, C, T or G). c. Checking for allele flipping and correcting allele flipping errors. d. Excluding strand-ambiguous SNPs. e. Excluding SNPs with a high missing genotype call rate in PD and SZ cohorts. (3) Filtering based on MAF: we dropped SNPs in the 1st and the 99th percentile of the distribution of differences in MAF to eliminate extreme outliers in the two output files. Finally, 4,070,160 SNPs remained to be used in the subsequent proxy-phenotype and prediction analyses.

### Proxy-phenotype method (PPM)

PPM [28, 29] was performed to identify shared novel genetic variants between PD and SZ. In the current study, SZ was used as the proxy phenotype. PPM is a two-stage approach. In the first stage, we applied a pre-specified p-value threshold of p < 1 × 10⁻⁵ to identify SZ-associated SNPs, consistent with previous studies [28, 29] and informed by evaluation of different cutoff thresholds. This was also to maximize the number of candidates forwarded to the next stage. A larger set of candidates increases the likelihood of capturing true positives. To select approximately independent SNPs from the SZ GWAS results, we performed clumping in PLINK v2.0 (https://www.cog-genomics.org/plink/2.0/postproc#clump) using r2 > 0.1 and 1,000 kb as the clumping parameters and the 1000 Genomes phase 3 (https://cncr.nl/research/magma/) European reference panel to estimate linkage disequilibrium (LD) among SNPs. This algorithm assigned the SNP with the smallest P value as the lead SNP in its clump. All SNPs in the vicinity of 1,000 kb around the lead SNPs that were correlated with it at r2 > 0.1 were assigned to this clump. The next clump was formed around the SNP with the next smallest P value, consisting of SNPs that have not been already assigned to the first clump. This process was iterated until no SNPs remained with P < 1 × 10^-5^. In the second stage, we firstly set 0.05 as p value threshold to filter out SNPs which were not significant for PD. Then we used Bonferroni correction setting a strict threshold (p < 5.92 × 10⁻⁵ (0.05/845)) to find more significant SNPs associated with PD.

To further validate the shared candidate SNPs identified through the two-stage PPM approach, we utilized a Bayesian statistical framework by conditioning on the SNPs’ effect sizes between PD and SZ. The analysis used a bivariate Bayesian regression model with residual correlations via the brms package v2.22.0 [35], incorporating effect sizes and standard errors as inputs. Weakly informative normal priors (N (0, 0.5)) were specified for intercepts, and a Lewandowski-Kurowicka-Joe (parameter 2) [36] prior was used for the residual correlation matrix. The model employed 4 chains with 4,000 iterations (2,000 warmup) and increased adaptation parameters (adapt_delta=0.99, max_treedepth=12) to ensure robust convergence. Following bonferroni multiple testing corrections, significantly associated SNPs across both disorders were identified and ranked by combined absolute effect sizes. Top 5 shared SNPs were listed in Supplementary table S1.

In order to investigate the novelty of the SNPs, LDassoc (https://ldlink.nih.gov/?tab=ldassoc) and LDproxy (https://ldlink.nih.gov/?tab=ldproxy) tools were utilized to determine whether these SNPs were in LD with the previously reported 90 PD-associated SNPs [21]. Additionally, we assessed LD between these SNPs and known variants reported from one previous study [25]. Thus, the SNPs related with previously reported ones were excluded. After this, we performed a look-up in the GWAS catalogue [37] (https://www.ebi.ac.uk/gwas/) to look at if these novel variants were reported in previous GWAS studies and associations with traits. In addition, regional plots were generated with the online tool LocusZoom to visualize the regional pattern of shared locus (https://my.locuszoom.org/gwas/upload/) [38].

### RAI1 rs11649804 exploration with scRNA-seq data from FOUNDIN-PD

*RAI1* rs11649804 genotype extraction: Induced pluripotent stem cells (iPSC) from patients of the Parkinson’s Progression Markers Initiative (PPMI) have been generated and characterized [30]. The PPMI iPSC lines were differentiated in vitro into dopaminergic neurons (DNs) and collected at day 65 using a protocol adapted from Kriks et al. [39] with minor modifications. Single-cell RNA sequencing indicated enrichment of *TH*, *MAP2*, and *SNCA* in neuronal cell types. Immunocytochemistry at day 65 showed that an average of 80% of cells (range: 52%–93%) were MAP2-positive neurons, while 20% (range: 4%–42%) were TH-positive. At the single-cell level, mature DNs were characterized by expression of *TH*, *ATP1A3*, *ZCCHC12*, *MAP2*, *SYT1*, and *SNAP25* (96,623 cells; 23% of total), whereas immature dopaminergic neurons (IDNs) expressed serotonergic markers *TPH1*, *SLC18A1*, *SLC18A2*, and *SNAP25* (41,267 cells; 10% of total). The cellsnp-lite [40] v1.2.3 tool in Mode 2b was used in pseudo-bulk manner to call genotypes in scRNA-seq BAMs using the whole genome sequencing WGS VCF file from chromosome 17. The cellsnp-lite analysis was performed primarily to determine *RAI1* genotypes and to stratify samples into CC, CA, and AA subgroups.

iPD, *GBA1*-PD and *LRRK2*-PD were categorized into three groups based on *RAI1* genotypes (iPD: N_CC_ = 12, N_CA_ = 14, N_AA_ = 3; *GBA1*-PD: N_CC_ = 6, N_CA_ = 6, N_AA_ = 4; *LRRK2*-PD: N_CC_ = 10, N_CA_ = 10, N_AA_ = 2). Due to the limited sample size in one of the *LRRK2*-PD genotype groups, differential expression analysis was not conducted for *LRRK2*-PD. Differential expression analysis among *RAI1* rs11649804 genotypes: The “FindAllMarkers” function in the Seurat v5 was used to identify differentially expressed genes (DEGs) across cell clusters in scRNA-seq data. On the other hand, genotypes in *RAI1* rs11649804 were added into metadata of sc-RNA seq data using the "AddMetaData" function in Seurat version v5. According to the mutation status, Seurat object was subsetted into two different subgroups with "subset" function: idiopathic PD without known pathogenic genetic variants and PD carrying *GBA1* N370S and E326K. Differential expression analysis between PD and controls was performed to evaluate *RAI1* expression, followed by separate differential gene expression analyses in iPD and *GBA1*-PD. Non-parametric Wilcoxon rank sum test was implemented in the “FindMarkers” function of Seurat v5 with default parameters (logfc.threshold = 0.1) between homozygous non-risk allele carriers and carriers of the *RAI1* rs11649804 risk allele (i.e., CA vs. CC, AA vs. CC and CA/AA vs. CC). The C allele was the disease-associated allele, whereas A was non-risk allele.

Based on cell types, scRNA-seq data in dopaminergic neurons (DNs) and immature dopaminergic neurons (IDNs) were extracted from each subset of the Seurat object. Using the "FetchData" function from the Seurat v5, we extracted normalized quantitative *RAI1* expression values in all DNs and IDNs. Then, basic functions in the dplyr package were used to calculate mean expression by averaging values across all DNs/IDNs per individual as well as stratify data by genotype (CC, CA, AA). In this regard, we got *RAI1* expression profiles across allelic dosages of *RAI1* rs11649804. Differential gene expression between pairwise comparisons of *RAI1* rs11649804 genotypes (e.g., CA vs. CC, AA vs. CC and CA/AA vs. CC) were assessed in DNs and IDNs.

Gene Ontology (GO) enrichment analysis and visualization: GO enrichment analysis for biological processes was performed using EnrichR [41] library GO_Biological_Processes_2025 (https://maayanlab.cloud/Enrichr/). The EnrichR combined score was used to rank the significance of GO terms, calculated by the logarithmic transformation of the p-value obtained from Fisher’s exact test, multiplied by the z-score representing the deviation from the expected rank. Differentially expressed genes (DEGs) were selected based on an adjusted p-value (p_val_adj < 0.05) and a minimum expression threshold (pct.1 >= 0.5, pct.2 >= 0.5) across the genotype comparisons (CA vs. CC, AA vs. CC, and CA/AA vs. CC). The top five most significant GO terms per comparison were selected based on the highest combined score. For visualization, heatmaps across the genotype comparisons (CA vs. CC, AA vs. CC, and CA/AA vs. CC) were generated using hierarchical clustering to group GO terms, implemented in R. Volcano plots were generated with " SCpubr 2.0.2" package in R.

### Cross-phenotype comparison of GWAS significance among neurodegenerative and psychiatric disorders

P-values for the *RAI1* variants (rs11649804 and rs12941356) were extracted from GWAS summary statistics of neurodegenerative diseases (PD [21], AD [42], FTD [43], ALS [44], MSA [45], PSP [46]), psychiatric disorders (SZ [20], ADHD [47], AN [48], ASD [49], BIP [50], MDD [51], OCD[52], PPD [53], PTSD [54]) and smoking initiation [55]) to compare the strength of statistical evidence for association across traits.

### Polygenic risk score (PRS) prediction in Tuebingen Parkinson cohort (TUEPAC)

The PD GWAS summary statistics was used for base data [21]. Imputed genotypes of 944 cases from Tuebingen, which were not included in the base data, were used for target data. Samples were primarily genotyped on Neurochip [56]. Quality control referred to one previous literature [57]. Polygenic risk scores(PRS) were calculated using PRSice-2 v2.3.5 (https://github.com/choishingwan/PRSice) [58, 59]. Linkage disequilibrium (LD) and clumping was performed using default settings (r2 = 0.1 and distance of 250kb). A p-value of 0.05 was chosen as the threshold to exclude non-significant SNPs based on our previous work [31]. PRS were standardized before analysis. Demographic information (sex and age at onset) was collected. And the Unified Parkinson’s Disease Rating Scale part III (UPDRS III) [60] and non-motor symptoms questionnaire (NMSQ) [61] were assessed. Polygenic risk scores (PRSs), including PD_all, PD_concordant, PD_discordant, and SZ_all, were constructed to predict clinical outcomes in the TUEPAC cohort. PD_all included all overlapping SNPs weighted by PD

GWAS effect sizes, whereas PD_concordant and PD_discordant included overlapping SNPs with concordant and discordant effect directions between PD and SZ, respectively, also weighted by PD GWAS effect sizes. SZ_all included all overlapping SNPs weighted by SZ GWAS effect sizes. After clumping and applying a P-value threshold of 0.05, 6,744, 3,689, 3,617, and 10,276 independent SNPs were retained for the PD_all, PD_concordant, PD_discordant and SZ_all, respectively. Associations between PRSs and age at onset (AAO) were assessed using multivariable linear regression adjusted for sex and the first four principal components (PCs). Associations between PRSs and clinical outcomes were also evaluated using linear regression adjusted for sex, age at visit, disease duration and the first four PCs.

### Cell-type association with genetic risk of PD

Association analysis of cell type-specific expressed genes with genetic risk of PD was performed as described previously [30], using Multi-marker Analysis of GenoMic Annotation (MAGMA) v1.10, in order to identify disease-relevant cell types in the data [62, 63]. MAGMA, as a gene set enrichment analysis method, was utilized to test cell type association with PD_concordant and PD_discordant SNP sets. Competitive gene set analysis was performed on SNP p-values from the large scale PD GWAS summary statistics including 23andMe data [21]. European samples from the 1000 Genomes Project Phase 3 were used as the reference panel to estimate linkage disequilibrium (LD) between SNPs. SNPs were mapped to genes using NCBI GRCh37 build. Gene boundaries were defined as the transcribed region of each gene. An extended window of 10 kb upstream and 1.5 kb downstream of each gene was added to the gene boundaries.

## Results

The complete analysis workflow was presented in Figure 1. After quality control using the MungeSumstats package and filtering out SNPs with extreme allele frequencies (0.01 < effect allele frequency < 0.99), 4,174,261 SNPs from the SZ GWAS and 4,318,815 SNPs from the PD GWAS were retained. Merging the two datasets by SNP ID yielded 4,070,160 overlapping SNPs for further analysis. Using the PPM approach, we first identified 845 SNPs associated with SZ at a threshold of P_SZ_ < 10^-5^. Of these, 123 SNPs showed nominal association with PD (P_PD_ < 0.05), and 14 SNPs remained significant after Bonferroni correction (P_PD_ < 0.05/845 = 5.92 × 10^−5^; Table 1), highlighting potential shared genetic risk between the two disorders. After applying LD-based filtering, eleven of the shared SNPs were excluded due to LD with 90 previously reported PD GWAS loci [21] or SNPs identified in prior cross-disorder studies, such as those by Smeland et al [25]. As a result, three shared novel SNPs-rs2240921(*ITIH3*), rs11649804(*RAI1*), and rs9607782(*EP300-AS1*) were identified (Table 1). These variants exhibit discordant effect directions in both SZ and PD, suggesting true pleiotropic associations rather than spurious overlap driven by LD with known risk loci.

**Figure 1.**
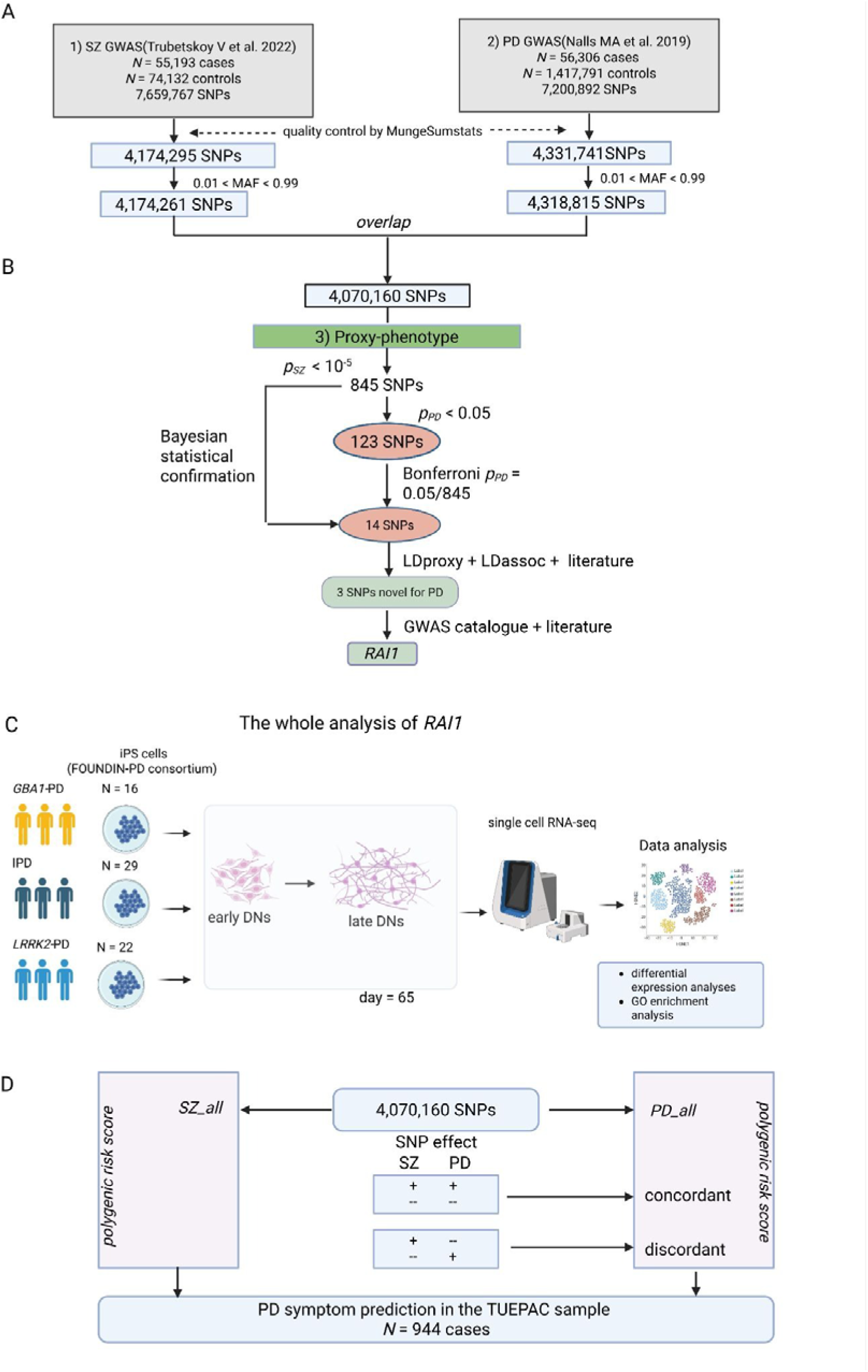
Workflow of the proxy-phenotype analyses. (A) Quality control and harmonization of Schizophrenia (SZ) and Parkinson’s disease (PD) GWAS datasets followed by identification of overlapping SNPs. (B) Proxy-phenotype analysis and Bayesian statistical prioritization of shared variants between SZ and PD highlighting *RAI1*. (C) Single-cell RNA-seq study design using iPSC-derived dopaminergic neurons from control, iPD, and *GBA1*-PD samples for downstream *RAI1*-focused analyses. (D) Polygenic risk score prediction of clinical outcomes in PD patients from the Tübingen Parkinson Cohort (TUEPAC). SZ and PD GWAS results are based on previously reported SZ and PD GWAS summary statistics. There was no sample overlap between the two studies. The TUEPAC data collection was not included in either the SZ or the PD meta-analysis. Proxy-phenotype analysis was performed using 4,070,160 autosomal SNPs that passed quality control. In addition, single-cell RNA sequencing data from FOUNDIN-PD were used to investigate the expression pattern of *RAI1* across CC/CA/AA genotype groups. The C allele is the risk-associated allele, whereas the A allele is protective. Abbreviations: IPD, Idiopathic Parkinson’s Disease.

**Table 1.** Overview of SNPs overlapping between Parkinson’s disease (PD) and schizophrenia (SZ) identified using the proxy-phenotype method (PPM).

|  |  |  |  |  |  |  |  |  |  |  |  |  |  |  |  |  |
| --- | --- | --- | --- | --- | --- | --- | --- | --- | --- | --- | --- | --- | --- | --- | --- | --- |
| SNP | variant consequence | mapped gene | PD GWAS |  |  |  | SZ GWAS |  |  |  | SZ GWAS without WTCCC |  | Direction of effect | LD assoc | LD proxy | Smeland et al. |
|  |  |  | EA | MAF | P | Beta | EA | MAF | P | Beta | P | Beta |  |  |  |  |
| **rs2240921**<br>**3:52830764:C:T** | synonymous | *ITIH3* | T | 0.12 | 2.37e-05 | 0.060 | C | 0.89 | 1.88e-06 | 0.064 | 5.56e-06 | 0.065 | dis | N | N |  |
| rs35225200<br>4:103146888:A:C | intergenic | *SLC39A8, BANK1* | A | 0.92 | 2.81e-06 | 0.089 | A | 0.91 | 8.75e-19 | -0.15 | 1.08e-16 | -0.14 | dis | N | N |  |
| rs9379968<br>6:27244590:G:A | intergenic | *POM121L2,CDCA7P1* | A | 0.48 | 4.00e-07 | 0.048 | G | 0.54 | 1.37e-11 | 0.058 | 2.69e-11 | 0.061 | dis | Y | Y |  |
| rs9379979<br>6:27299614:C:T | intergenic | *ZNF391* | T | 0.059 | 3.23e-5 | -0.09 | C | 0.93 | 4.06e-6 | -0.081 | 9.40e-05 | -0.072 | dis | Y | Y |  |
| rs2734335<br>6:31893944:G:A | intronic | *C2* | A | 0.51 | 1.07e-5 | -0.042 | G | 0.48 | 7.93e-9 | -0.05 | 2.98e-08 | -0.051 | dis | Y | Y |  |
| rs9270836<br>6:32570325:A:G | intergenic | *HLA-DRB5* | A | 0.28 | 1.18e-7 | 0.07 | A | 0.33 | 1.743e-8 | 0.068 | 6.08e-07 | 0.064 | con | Y | Y |  |
| rs298602<br>8:130932174:C:T | intronic | *CYRIB* | T | 0.49 | 2.14e-08 | 0.052 | C | 0.54 | 4.15e-07 | 0.044 | 9.83e-07 | 0.045 | dis | Y | Y |  |
| rs546381<br>11:133760651:T:G | intergenic | *IGSF9B* | T | 0.56 | 4.83e-07 | 0.047 | T | 0.55 | 3.77e-07 | 0.044 | 1.24e-07 | 0.048 | con | Y | Y |  |
| rs17128077<br>14:55386034:C:T | intergenic | *GCH1* | T | 0.14 | 9.31e-06 | -0.061 | C | 0.85 | 1.36e-06 | -0.059 | 3.40e-06 | -0.060 | dis | Y | Y |  |
| rs1861759<br>16:50745583:T:G | synonymous | *NOD2* | T | 0.61 | 1.78e-08 | 0.055 | T | 0.61 | 2.72e-06 | 0.041 | 3.76e-07 | 0.047 | con | Y | Y |  |
| **rs11649804**<br>**17:17696755:C:A** | missense | *RAI1* | A | 0.30 | 8.49e-7 | -0.051 | C | 0.69 | 6.66e-7 | -0.047 | 4.41e-07 | -0.050 | dis | N | N |  |
| rs117499775<br>17:44078618:T:C | intronic | *MAPT* | T | 0.97 | 1.11e-05 | 0.18 | T | 0.97 | 8.52e-06 | 0.14 | 2.73e-05 | 0.14 | con | Y | Y |  |
| rs199497<br>17:44866602:T:C | intronic | *WNT3* | T | 0.83 | 3.32e-17 | 0.12 | T | 0.85 | 2.79e-06 | 0.059 | 4.08e-05 | 0.054 | con | Y | Y |  |
| **rs9607782**<br>**22:41587556:T:A** | intronic | *EP300-AS1* | A | 0.24 | 2.96e-5 | -0.058 | T | 0.74 | 4.97e-14 | -0.076 | 3.08e-12 | -0.074 | dis | N | N |  |

**913** **914** Notes: The effect allele (EA) denoted the allele for which the effect size was estimated in PD and SZ genome wide **915** association study (GWAS) summary statistics. Therefore, the same SNP may have different reported effect alleles **916** (EA) across datasets. Beta values and minor allele frequency (MAF) were presented relative to the EA shown for **917** each GWAS and the concordance of effect directions should be evaluated after allele harmonization across **918** datasets. LD, Linkage disequilibrium; N means no, there were not any other SNPs in PD GWAS and Smeland et al. **919** associated with one of these fourteen SNPs; Y means yes, vice versa. con, concordant, dis, discordant. SNPs shown **920** in bold indicate novel variants shared between PD and SZ that were not in linkage disequilibrium with any **921** previously reported SNPs from PD GWAS or other related studies. Coordinates are based on human genome **922** assembly GRCh37 build. **923** **924** **925** **926** **927** **928** **929** **930** **931** **932** **933** **934** **935**

We focused on rs11649804, located in the *RAI1* gene, as it represents a missense variant in the coding region of the gene. Single-cell RNA sequencing from Foundin-PD was used in this analysis, as described in the methods and in previous work [30]. Briefly, at the single-cell level, mature dopaminergic neurons (DNs) were defined by expression of *TH, ATP1A3, ZCCHC12, MAP2, SYT1,* and *SNAP25*, whereas immature dopaminergic neurons (IDNs) expressed serotonergic markers *TPH1, SLC18A1, SLC18A2,* and *SNAP25*. *RAI1* expression was significantly reduced in iPD iPSC derived dopaminergic neurons (DNs) (*N_iPD_* = 29 vs *N_HC_* = 9, adjusted p = 1.34e-10, avg_log2FC = -0.17) and immature dopaminergic neurons (IDNs) (*N_iPD_* = 29 vs *N_HC_* = 9, adjusted p = 9.30e-38, avg_log2FC = -0.51) compared with healthy controls. Similarly, decreased *RAI1* expression was observed in IDNs from *GBA1*-PD (*N_GBA1-PD_* = 16 vs *N_HC_* = 9, adjusted p = 3.09e-06, avg_log2FC = -0.33) and *LRRK2*-PD (*N_LRRK2-PD_* = 22 vs *N_HC_* = 9, adjusted p = 1.93e-06, avg_log2FC = -0.16) compared with controls (Figure S1). Given that one of the genotype groups in the control and *LRRK2*-PD cohorts had less than three samples, genotype-based *RAI1* expression comparisons (CC/CA/AA) were performed only within the iPD and *GBA1*-PD groups. In iPD DNs, *RAI1* expression was significantly increased in homozygous risk SNP carriers compared to homozygous non-risk SNP carriers (*N_CC_*= 12 vs *N_AA_*= 3, adjusted p = 0.0033, avg_log2FC = 0.12). In contrast, in *GBA1*-PD IDNs, we observed a lower *RAI1* expression in risk SNP carriers compared to non-risk SNP carriers (*N_CC_* = 6 vs *N_AA_* = 4, adjusted p = 3.88e-08, avg_log2FC = -0.33; *N_CC_* = 6 vs *N_CA/AA_* = 10, adjusted p = 9.47e-07, avg_log2FC = -0.22) subsets in IDNs (Figure S2). The distribution of up- and down- regulated genes between the disease associated SNP genotype (CC) and control SNP genotype (CA/AA) was shown in Figure 2. Differential expression analysis revealed significantly reduced *RBP1* expression and increased *ENO1*, *PGK1*, and *PKM* expression in *GBA1*-PD patients carrying the risk-associated C allele compared to those homozygous for the protective A allele (Figure 2B).

**Figure 2.**
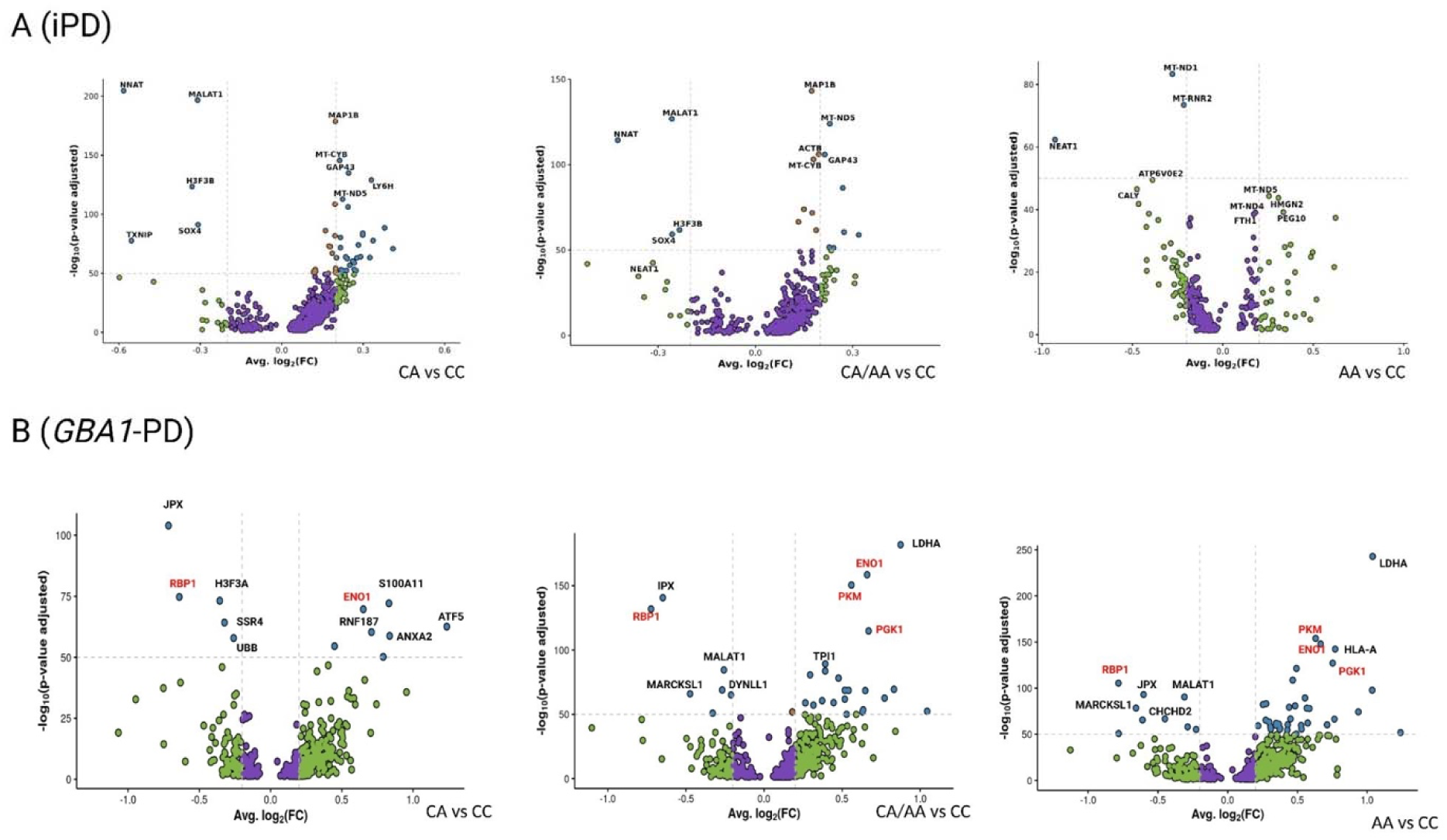
Volcano plot for up- and down- regulated genes by *RAI1* rs11649804 genotype. (A) Differentially expressed genes of *RAI1* rs11649804 in idiopathic PD dopaminergic neurons; (B) Differentially expressed genes of *RAI1* rs11649804 in *GBA1* immature dopaminergic neurons. The C allele is the risk-associated allele, whereas the A allele is protective.

To investigate biological pathways influenced by *RAI1* expression, we performed GO enrichment analysis on differentially expressed genes in iPSC-derived DNs and IDNs from iPD and *GBA1*-PD individuals. Differentially expressed markers and enriched GO terms across *RAI1* genotypes in immature dopaminergic neurons and dopaminergic neurons in iPD and *GBA1*-PD patients were summarized in Table S2–S5. In iPSC derived DNs from iPD patients, differential gene expression analysis between the control SNP genotype (AA) and the disease associated SNP genotype (CC) groups identified a total of 264 significant genes (adjusted p < 0.05 & pct.1 >= 0.5 & pct.2 >= 0.5) down-regulated, whereas 76 significant genes were up-regulated. GO enrichment analysis revealed that down-regulated genes in DNs were associated with Aerobic Electron Transport Chain (GO:0019646), Mitochondrial Electron Transport, Cytochrome C to Oxygen (GO:0006123), Mitochondrial ATP Synthesis Coupled Electron Transport (GO:0042775), Cellular Respiration (GO:0045333), and Plus-End-Directed Organelle Transport Along Microtubule (GO: 0072386) terms. In addition, differential gene expression analysis between CA/AA and CC groups identified a total of 430 significant genes up-regulated. GO enrichment analysis showed that up-regulated genes in DNs were associated with Substantia Nigra Development (GO:0021762) (Figure 3). In iPSC derived IDNs from *GBA1*-PD patients, differential gene expression analysis between CA and CC groups identified a total of 365 significant genes up-regulated, while 166 significant genes were down-regulated. GO enrichment analysis revealed that down-regulated genes in IDNs were associated with Mitochondrial Electron Transport, NADH to Ubiquinone (GO: 0006120), Cellular Respiration (GO:0045333), Aerobic Electron Transport Chain (GO:0019646), Mitochondrial ATP Synthesis Coupled Electron Transport (GO:0042775), Proton Motive Force-Driven Mitochondrial ATP Synthesis (GO: 0042776) and Oxidative Phosphorylation (GO:0006119) terms (Figure 3).

**Figure 3.**
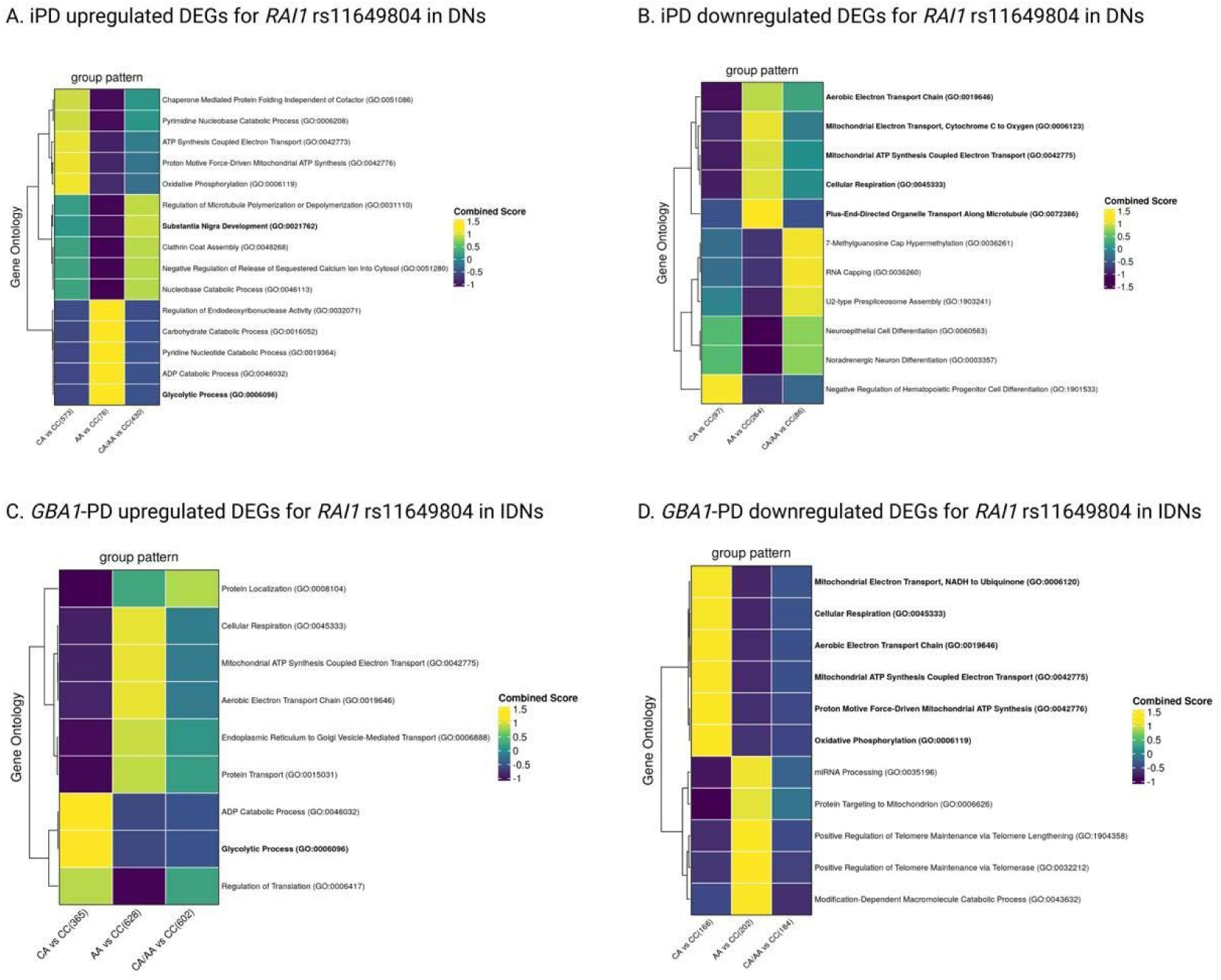
Gene ontology (GO) and enrichment analysis of *RAI1* rs11649804-linked differentially expressed genes in dopaminergic neurons (DNs) and immature dopaminergic neurons (IDNs).

To further visualize the regional association signals and LD structure of the shared loci, LocusZoom plots were generated for each locus (Figure 4A-4C; Figure S3-S6). As 2025 PD GWAS was not available at the time of initial analysis, LocusZoom plots were generated using both the 2019 and 2025 PD GWAS datasets. Comparison of these plots showed stronger association signals at the *RAI1* locus in the 2025 PD GWAS [64] than in the 2019 PD GWAS [21], including rs12941356 (p = 1.09 × 10^-11^ vs. 1.31 × 10^-7^) and rs11649804 (p = 5.17 × 10^-7^ vs. 8.49 × 10^-7^). In the 2025 PD GWAS, rs12941356 reached genome-wide significance and emerged as the lead SNP at the *RAI1* locus, whereas rs11649804 was the lead SNP in SZ, as shown in Figure 4. These results suggest that SZ GWAS might help to identify variants that later reach genome-wide significance in larger PD GWAS. Additionally, as shown in Figure 4D, rs12941356 and rs11649804 exhibited phenotype-specific association signals, with rs12941356 showing a strong association with PD, and rs11649804 with SZ, while remaining largely nonsignificant across other neurodegenerative and psychiatric phenotypes. The effects of two variants between neurodegenerative and psychiatric disorders, rs11649804 and rs12941356, were provided in Table S6. In addition, the shared genetic variants between PD and SZ and their previously reported GWAS Catalog associations were summarized in Table S7.

**Figure 4.**
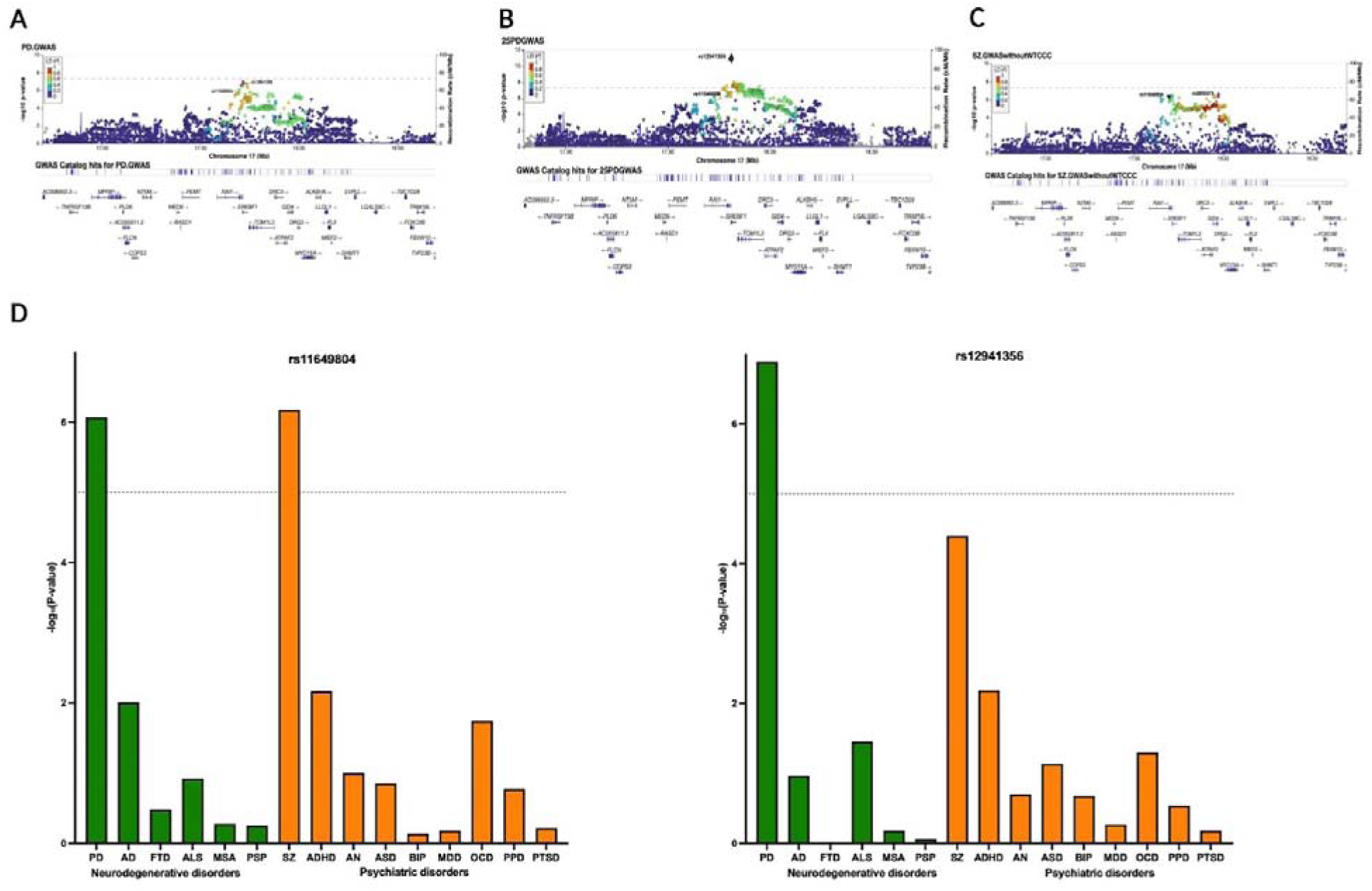
(A) rs11649804(17:17696755) in 2019 PD GWAS[21]. The Genome build is GRCh37; (B) rs11649804(17:17793441) in 2025 PD GWAS [54]. The Genome build is GRCh38; (C) rs11649804(17:17696755) in 2022 SZ GWAS[20] without WTCCC2 controls. The Genome build is GRCh37; (D) Comparative Analysis of GWAS P-values Across Neurodegenerative and Psychiatric Disorders. Bar plots showed the –log10 (p-value) from genome-wide association studies (GWAS) for two genetic variants: rs11649804 (left) and rs12941356 (right). Each bar represents a disorder, grouped by disease category: neurodegenerative disorders (green) and psychiatric disorders (orange). The y-axis represents the statistical significance (–log10 of p-value) of association with each disorder. A dashed horizontal line marks the threshold of p = 1e-5. Disorders included are: (1) Neurodegenerative: Parkinson’s disease (PD), Alzheimer’s disease (AD), Frontotemporal dementia (FTD), Amyotrophic lateral sclerosis (ALS), Multiple system atrophy (MSA), Progressive supranuclear palsy (PSP). (2) Psychiatric: Schizophrenia (SZ), Attention-deficit/hyperactivity disorder (ADHD), Anorexia nervosa (AN), Autism spectrum disorder (ASD), Bipolar disorder (BP), Major depressive disorder (MDD), Obsessive-compulsive disorder (OCD), Post-partum depression (PPD) and Post-traumatic stress disorder (PTSD).

To investigate the clinical relevance of shared genetic architecture between PD and SZ, we stratified overlapping variants based on the direction of their effects. We hypothesized that shared variants would differentially influence non-motor symptom burden in PD depending on effect direction. To test this, we constructed polygenic risk scores (PRS) based on concordant and discordant SNP sets and evaluated their association with clinical outcomes in the Tuebingen Parkinson cohort (TUEPAC). In an initial model including PD_all and SZ_all, PD_all significantly predicted both age of onset (beta = -0.07, P = 0.038) and non-motor symptom burden (beta = -0.16, P = 0.041), while SZ_all was not significant for either outcome (Table 2). To further dissect the contribution of shared genetic architecture, we stratified PD_all into concordant and discordant components and refit the model including PD_concordant, PD_discordant, and SZ_all. In this stratified model, only the discordant PRS remained a significant predictor of NMSQ (beta = -0.17, P = 0.034), indicating that the non-motor symptom association observed in the combined model is specifically attributable to variants with opposing effects across PD and SZ (Table 2). For age of onset, neither concordant nor discordant PRS reached significance after stratification (PD_concordant: beta = 0.041, P = 0.23; PD_discordant: beta = 0.044, P = 0.21), though both retained negative beta coefficients consistent with the direction observed in the combined model, suggesting that the age of onset signal was distributed across both directional components rather than localized to either alone (Table 2). SZ_all remained non-significant across all models and outcomes.

**Table 2.** Polygenic risk score prediction of age at onset and non-motor symptoms scales in the Tuebingen Parkinson Cohort sample.

|  |  | Age at onset<sup>a</sup> | UPDRS-III<sup>b</sup> | NMSQ<sup>b</sup> |
| --- | --- | --- | --- | --- |
| Baseline model |  |  |  |  |
| PD_all | Standardized beta | -0.070 | 0.040 | -0.16 |
|  | P value | 0.038* | 0.35 | 0.041* |
| SZ_all | Standardized beta | 0.024 | 0.021 | -0.031 |
|  | P value | 0.47 | 0.62 | 0.69 |
|  | Adj. R<sup>2</sup> | 0.018 | 0.068 | 0.022 |
| Model | P value | 0.002 | 9.28e-07 | 0.17 |
| Split model |  |  |  |  |
| Concordant | Standardized beta | -0.041 | 0.021 | -0.046 |
|  | P value | 0.23 | 0.63 | 0.55 |
| Discordant | Standardized beta | -0.044 | 4.21e-04 | -0.17 |
|  | P value | 0.21 | 0.99 | 0.034* |
| SZ_all | Standardized beta | 0.024 | 0.015 | -0.061 |
|  | P value | 0.50 | 0.74 | 0.45 |
|  | Adj. R<sup>2</sup> | 0.016 | 0.065 | 0.024 |
| Model | P value | 0.004 | 2.91e-06 | 0.17 |
| ΔR<sup>2</sup>(Split-baseline model) |  | -0.002 | -0.003 | 0.002 |
| n |  | 887 | 527 | 177 |

Notes: Linear regression, <sup>a</sup>Sex and the first four genetic principal components (PCs 1-4) were included as covariate. <sup>b</sup>Sex, age at visit, disease duration and PCs 1-4 were included as covariate. SE, standard error; UPDRS-III, Unified Parkinson's Disease Rating Scale part III; NMSQ, Non-Motor Symptoms Questionnaire.

To identify the cell types underlying the shared genetic architecture between PD and SZ, we performed MAGMA gene-set enrichment analysis using brain postmortem single-cell RNA sequencing data. Concordant variants were significantly enriched in oligodendrocytes (ODCs; P = 0.038), oligodendrocyte precursor cells (OPCs; P = 0.011), and microglia (MG; P = 0.026), implicating the full oligodendrocyte differentiation axis alongside neuroinflammatory processes in the shared genetic liability between the two disorders (Figure 5A). Discordant variants were enriched in ODCs (P = 0.018), MG (P = 0.025), and vascular cells (P = 0.023) (Figure 5B). Notably, while ODCs and MG were enriched in both SNP sets, OPCs were uniquely enriched in the concordant set and vascular cells were uniquely enriched in the discordant set, highlighting distinct cellular mechanisms underlying each directional component of the shared genetic architecture. As a sanity check, we repeated the MAGMA analysis excluding *SNCA*, *GBA1*, and *LRRK2*, and obtained similar results (Figure S7).

**Figure 5.**
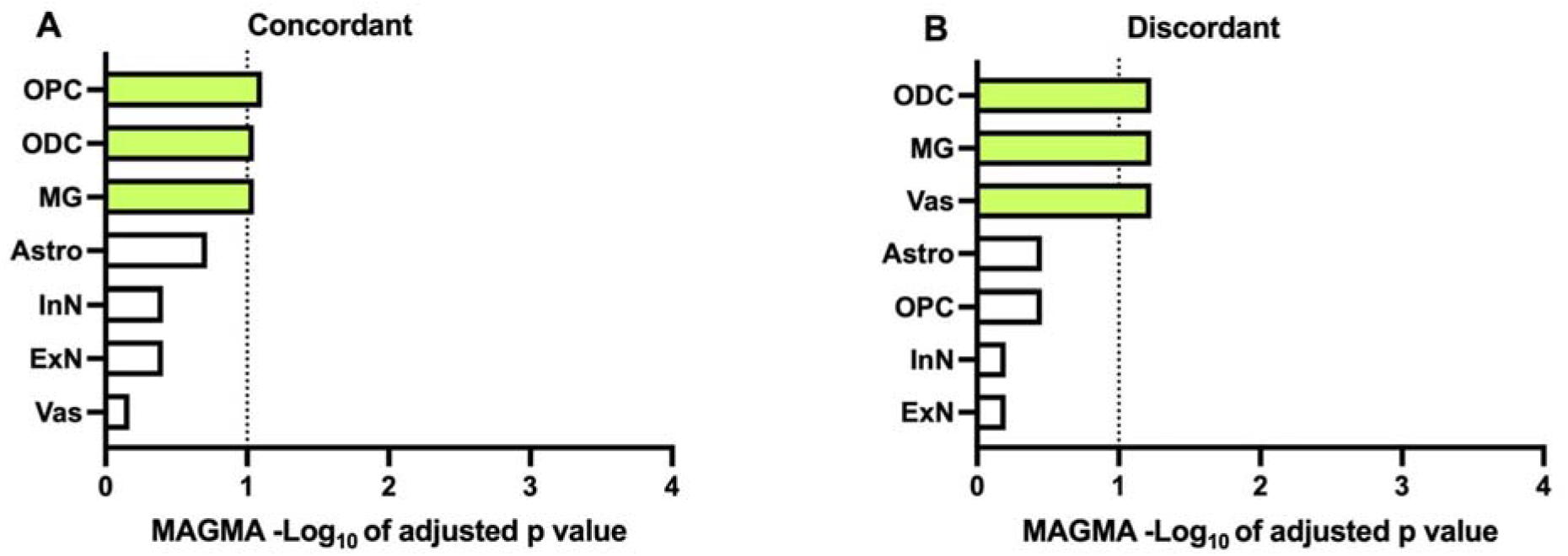
(A) Multi-marker analysis of genomic annotation (MAGMA) gene set enrichment based on PDconcordant SNP list in postmortem brain issue derived single cell RNA sequencing dataset [31].(B) MAGMA gene set enrichment based on PDdiscordant SNP list n postmortem brain issue derived single cell RNA sequencing dataset [31]. The dotted line indicates the FDR-adjusted significance threshold of 0.1. Abbreviations of various cell types: excitatory neurons (ExN), inhibitory neurons (InN), oligodendrocytes (ODCs), oligodendrocyte precursor cells (OPCs), microglia (MG), astrocytes (Astro) and vascular cells (Vas).

## Discussion

In the current study, we provide new insights into the shared genetic background between PD and SZ using non-overlapping GWAS summary statistics. First, the proxy-phenotype method (PPM) [28, 29] combined with Bayesian statistical framework were performed to identify fourteen shared SNPs, including three novel SNPs (one synonymous SNP, one intronic SNP and one missense SNP) and eleven SNPs which were in LD with previous reported SNPs (one synonymous SNP, six intergenic SNPs and four intronic SNPs). The missense SNP rs11649804, which results in a p.Pro165Thr amino acid substitution, was reported to be associated with PD risk [21, 64]. Subsequently, using single cell RNA-seq data from FOUNDIN-PD, we found *RAI1* rs11649804 risk allele was associated with increased expression in DNs of iPD patients and decreased expression in IDNs of *GBA1*-PD carriers, respectively. The changes in *RAI1* expression in DNs of iPD patients and in IDNs of *GBA1*-PD carriers resulted in enrichment of GO terms related with mitochondrial dysfunction.

Although the Brainstorm Consortium reported no genetic correlation between SZ and PD [65], a recent study identified nine SNPs (five with concordant and four with discordant effect directions) jointly associated with both PD and SZ using a conjunctional false discovery rate approach applied to GWAS summary statistics [25]. Consistent with these findings, our study identified fourteen SNPs (five concordant and nine discordant in effect direction), in line with the results reported in Smeland’s work [25]. One possible explanation is that genetic correlation analyses can only capture consistent effect directional SNPs and there is a mixed pattern of effect directions among overlapping SNPs. Thus, common variants shared between PD and SZ might contribute to overlapping biological mechanisms despite the absence of a global genetic correlation.

Overall, the majority of novel shared loci indicated a potential role of their related mechanisms, shared inflammatory pathophysiology between PD and SZ. Inter-alpha (globulin) inhibitor 3 (*ITIH3*), one of structurally related plasma serine protease inhibitors family (*ITIH1-5*), has been reported to be involved in inflammatory diseases, including rheumatoid arthritis and inflammatory bowel diseases [66]. Besides, multiple studies have confirmed the association of *ITIH3* with SZ [67–69]. The SNPs in E1A binding protein p300 (*EP300*) were found to be genetic risk factors in PD of multiple-ancestry populations [70] as well as in SZ [71, 72]. Furthermore, *EP300* was involved in the pathogenesis of several inflammatory conditions, including inflammatory bowel disease [73], pulmonary fibrosis [74], coronary artery disease [75].

*RAI1* is a major gene involved in Smith-Magenis syndrome (SMS), a neurological disorder typically caused by deletions at chromosome 17p11.2. Previous studies, including a clinical analysis of 36 patients with SMS-like features [76] and a reported case of co-occurring SMS and early-onset PD [77], have identified several non-motor symptoms that overlap with those observed in PD and SZ, such as cognitive impairment, sleep disturbances and anxiety. GWAS have also implicated the associations of *RAI1* with PD risk [78, 79]. In addition, our results showed reduced *RAI1* expression in PD compared with controls, consistent with *RAI1* expression levels previously reported in individuals of SMS [76].

In the current study, single cell RNA-seq data was utilized to investigate genotype-specific differences in *RAI1* expression, as well as to identify the biological pathways associated with changes in *RAI1* expression. Retinol Binding Protein 1 (*RBP1*) is an intracellular chaperone that binds retinol and retinal with high affinity and thereby facilitates retinoic acid (RA) biosynthesis [80]. In addition, RA production was reported to be influenced directly by *RBP1* expression [81]. Retinoic acid induced-1 (*RAI1*) is a dosage-sensitive gene and *RAI1* mRNA expression was regulated via disrupting the binding of Retinoid X Receptor-Retinoic acid receptor (RXR-RAR) [82]. Transcriptomic analysis showed reduced *RAI1* expression in *GBA1*-PD patients, while the volcano plot further supported this observation, with reduced RBP1 expression in the disease associated SNP genotype of *RAI1*. Together, these findings suggested that altered expression of both *RAI1* and *RBP1* genes may reflect disruption of RA-related pathways in *GBA1*-PD. Conversely, *ENO1*, *PGK1* and *PKM*, all of which were involved in glycolysis, were upregulated in *GBA1*-PD patients. In line with this observation, GO analysis showed enrichment of upregulated genes associated with a glycolytic phenotype in iPD and *GBA1*-PD groups. This may reflect a compensatory cellular response in which cells increase glycolytic activity to offset energy deficiency arising from mitochondrial dysfunction in PD patients carrying the disease-associated SNP genotype. Our study uncovered *RAI1*-associated mechanisms in PD at transcriptional level and suggested a potential shared biological pathway mediated by *RAI1* dysregulation between PD and SZ. A previous study reported impaired serotonin metabolism in *RAI1*-transgenic mice, suggesting that *RAI1* was involved in serotonin pathway regulation in a dosage-dependent manner [83]. In the current study, the serotonergic marker *TPH1* was expressed in iPSC-derived IDNs, and *RAI1* expression distinguished *GBA1*-PD from controls specifically in this cell population. Together, these findings suggested that serotonergic dysregulation may represent a potential shared mechanism to both PD and SZ.

Notably, when comparing LocusZoom plots from the 2019 [21] and 2025 [64] PD GWAS, we observed that the association signal at the *RAI1* locus was strengthened in the 2025 study, with the lead SNP surpassing genome-wide significance threshold. The PPM method implemented in our study may therefore facilitate the discovery of novel PD susceptibility loci in future GWAS with increased sample size. Furthermore, the shared genetic signal at the *RAI1* locus implicated involvement of common biological pathways contributing *to the* etiology of both disorders. Our finding therefore raised the possibility that genetic variants at the *RAI1* locus might contribute to both motor and non-motor symptoms observed in PD and SZ. Future studies leveraging longitudinal phenotyping in other larger cohorts will be essential to validate association between *RAI1* and non-motor symptoms in PD. On the other hand, rs12941356 and rs11649804, the lead SNPs within *RAI1* locus in PD and SZ respectively, showing minimal pleiotropic effects across other psychiatric related disorders. This pattern of association suggested that these variants may represent disorder-specific genetic signals uniquely shared between PD and SZ, as supported by the PPM analysis.

The stratification of PD genetic liability into concordant and discordant components revealed a clinically meaningful dissociation between disease outcomes. The non-motor symptom association localized specifically to the discordant PRS, suggesting that variants with opposing effects across PD and SZ capture biological pathways distinctly relevant to the non-motor dimensions of PD, independent of broader PD genetic burden. Cell type enrichment analysis using brain postmortem single-cell data revealed partially overlapping but distinct cellular profiles for concordant and discordant SNP sets, providing important biological context for the clinical associations observed. Both sets were enriched in mature oligodendrocytes and microglia, suggesting that oligodendrocyte lineage integrity and neuroinflammatory processes represent shared features of the genetic architecture linking PD and SZ regardless of effect direction. Beyond this shared signal, concordant variants were additionally enriched in OPCs, implicating the full oligodendrocyte differentiation axis. In contrast to concordant variants, discordant variants showed enrichment in vascular cells. The discordant genetic architecture may confer a vascular component of PD that partially recapitulates the non-motor symptom profile of vascular parkinsonism [84]. Together, these findings raise the possibility that discordant genetic architecture contributes to a more genetically driven, relatively reduced non-motor involvement. Further work dissecting specific non-motor symptom domains in a larger cohort, particularly psychiatric and cognitive subdomains given their phenotypic overlap with SZ, will be necessary to clarify whether the observed association reflects a buffering of neuropsychiatric features or a broader effect on non-motor symptom progression.

Several limitations should be acknowledged. First, although the effect of *RAI1* observed in this study was a little weak, our analyses relied primarily on transcriptomic and bioinformatic approaches without functional validation. Therefore, these findings should be interpreted with caution, and further experimental studies will be essential to confirm the observed transcriptomic changes in iPSC-derived dopaminergic neurons or human postmortem brain tissue. Second, our analyses were restricted to European-ancestry GWAS datasets due to limited sample sizes in other populations, which may affect the generalizability of the findings. In addition, phenotypic heterogeneity within the SZ GWAS may influence the observed genetic overlap, as GWAS signals could reflect a mixture of partially distinct biological subtypes, thereby complicating the interpretation of the genetic relationship with PD. Furthermore, some shared SNPs may influence PD and SZ through independent or only partially overlapping pathways. Therefore, these findings should be interpreted as evidence of shared genetic overlap rather than a shared causal mechanism. In terms of PPM, this approach represents a screening strategy that depends on the assumption that the proxy phenotype (SZ) is informative for the target trait (PD). Its performance is also sensitive to the pre-specified threshold for selecting candidate SNPs, and the optimal trade-off is hard to determine in practice. In the current study, we selected the optimized threshold based on prior literature [28, 29] and power calculations.

In addition, the association between the PD-discordant SNP set and PRS was evaluated in a relatively limited sample size; therefore, validation in larger independent cohorts is needed.

Overall, this study leveraged large-scale GWAS summary statistics to investigate the shared genetic architecture between PD and SZ, identifying three novel SNPs exhibiting discordant directional effects across both disorders. By integrating transcriptomic analyses, we further explored the functional relevance of these loci, implicating inflammatory pathways and mitochondrial dysfunction as key convergent mechanisms. These findings suggest that targeting shared molecular pathways may offer promising therapeutic strategies for individuals affected by neurological and psychiatric disorders. In addition, we identified, for the first time, a subset of SNPs discordant between PD and SZ, were enriched in vascular cell types, and had a polygenic burden associated with non-motor manifestations of PD. Future efforts involving large, deeply phenotyped cohorts and experimental studies will be critical to elucidate the biological significance of these shared genetic risk factors and their contributions to disease subtypes.

## Supporting information

Cohort overview of PD and SZ GWAS

## Data Availability

The SZ GWAS summary statistics are available on the website, https://www.med.unc.edu/pgc/download-results/scz/. The PD GWAS summary statistics from 23andMe (including data from Chang et al. 2017 and Nalls et al. 2014) are available to qualified researchers through 23andMe. Access requires an agreement to protect participant privacy. For details and to apply, visit: research.23andme.com/collaborate/#publication. The single cell RNA sequencing data are available on the website https://www.foundinpd.org/.

https://research.23andme.com/collaborate/#publication

https://www.foundinpd.org/

## Supplementary material

### Supplementary tables and figures

Supplementary Document 1: Cohorts overview of PD GWAS and SZ GWAS.

Table S2 (iPD_DE_markers): Marker list with differential analysis in iPD subset.

Table S3 (IPD_GO_terms): GO terms list for up/down-regulated genes in CC/CA/AA comparisons in IDNs and DNs in IPD subset.

Table S4 (GBA1_DE_markers): Marker list with differential analysis in *GBA1* subset.

Table S5 (GBA1_GO_terms): GO terms list for up/down-regulated genes in CC/CA/AA comparisons in IDNs and DNs in *GBA1*-PD subset.

## Author Contributions

V.B. conceived the project. T.G. and V.B. provided the resources and supervised the study. M.S. contributed the genotype data for the Tübingen Parkinson Cohort. I.W., B.R., R.K., and K.B. collected the clinical data for the Tübingen Parkinson Cohort. W.S. performed the whole analysis, data visualization and drafted the manuscript. N.K. contributed to the single-cell analysis. A.B. and F.S. analyzed the GWAS data from psychiatric disorders. V.B. and M.D. supervised the data analysis. M.D., M.M., V.B. and T.G. contributed to the guidance of the whole process and manuscript check. W.S., M.M. and V.B. wrote the manuscript. All authors contributed to and reviewed the manuscript.

## Acknowledgments

We would like to thank 23andMe Research Team, System Genomics of Parkinson’s Disease (SGPD) Consortium, the International Parkinson’s Disease Genomics Consortium, and Schizophrenia Working Group of the Psychiatric Genomics Consortium to provide PD and SZ GWAS summary statistics. The single cell RNA sequencing data used in this article was obtained from the PPMI database (www.ppmi-info.org/data) and was supported by Michael J. Fox Foundation for Parkinson’s Research. We also thank the COURAGE-PD Consortium for providing genotype data from the Tübingen Parkinson Cohort. The Courage-PD consortium is supported by the EU Joint Program for Neurodegenerative Disease research (JPND https://neurodegenerationresearch.eu).

## Code availability

Source code for the whole analyses is available at https://github.com/wenhuasun/PD-SZ/tree/main

