## Supplementary Figures and Tables for "Unraveling the Genetic Overlap Between Parkinson’s Disease and Schizophrenia Through Genome-wide Association and Cell-Type Specific Transcriptomic Analysis"

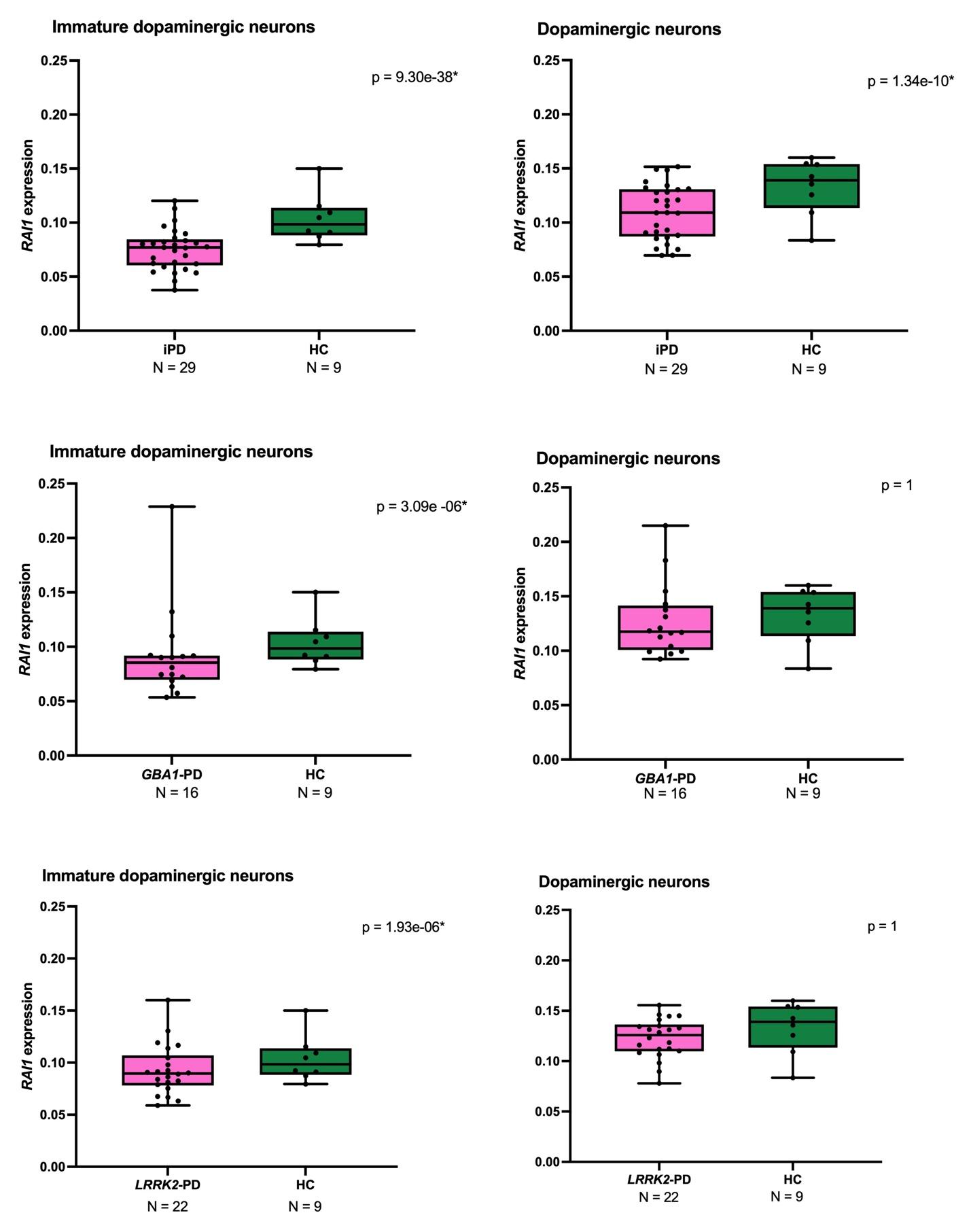

Figure S1. Expression of *RAI1* rs11649804 in dopaminergic neurons and immature dopaminergic neurons at the single-cell level comparing idiopathic Parkinson’s disease (iPD), *GBA1*-PD, and healthy controls (HC).

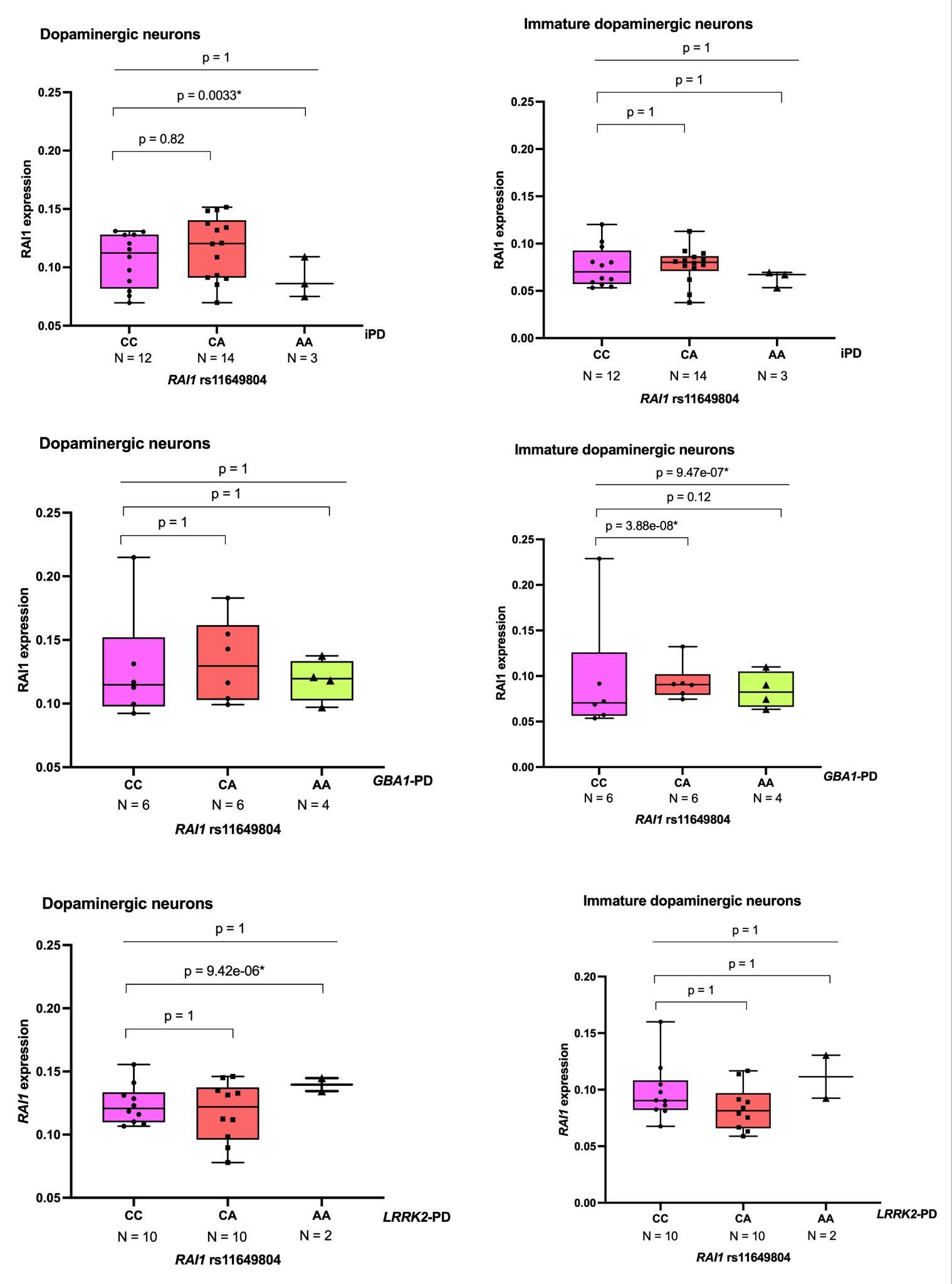

Figure S2. Expression of *RAI1* rs11649804 in dopaminergic neurons and immature dopaminergic neurons at the single-cell level between idiopathic Parkinson's disease (iPD) and *GBA1*-PD. The C allele is the risk-associated allele, whereas the A allele is protective.

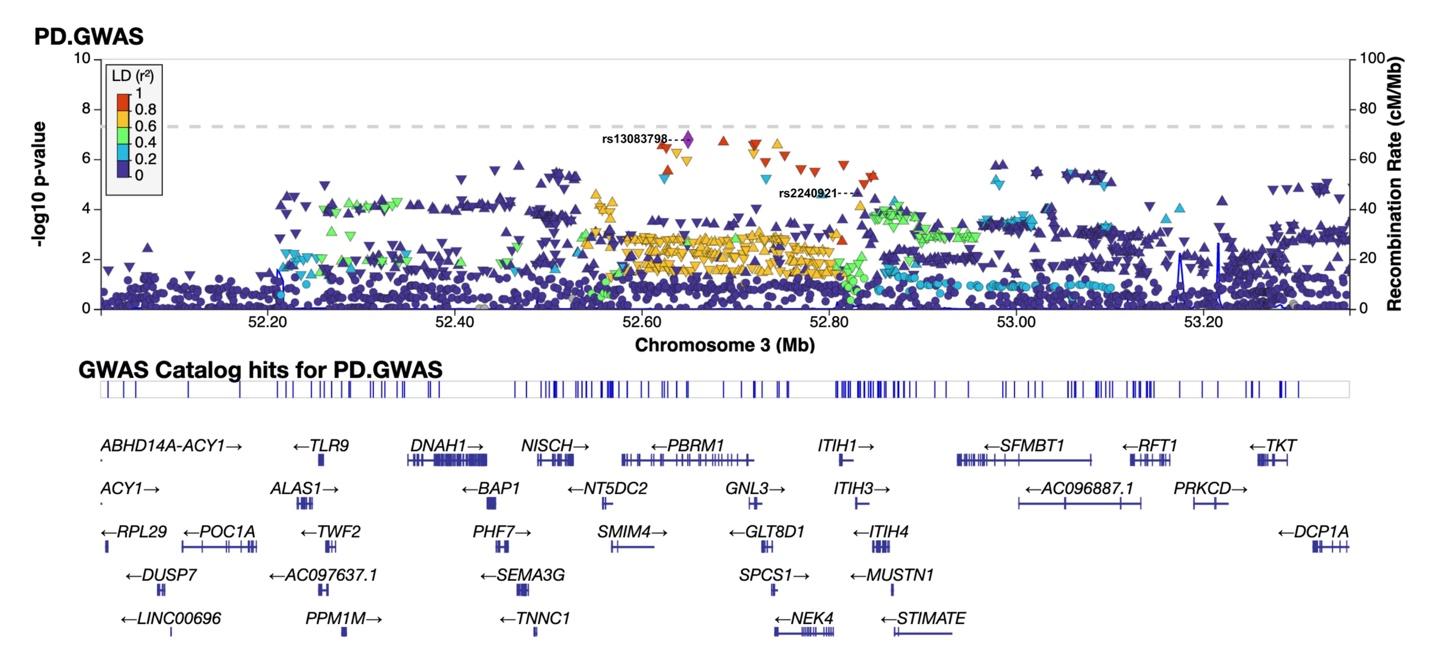

Figure S3a. rs2240921(3:52830764) in 2019 PD GWAS. The Genome build is GRCh37.

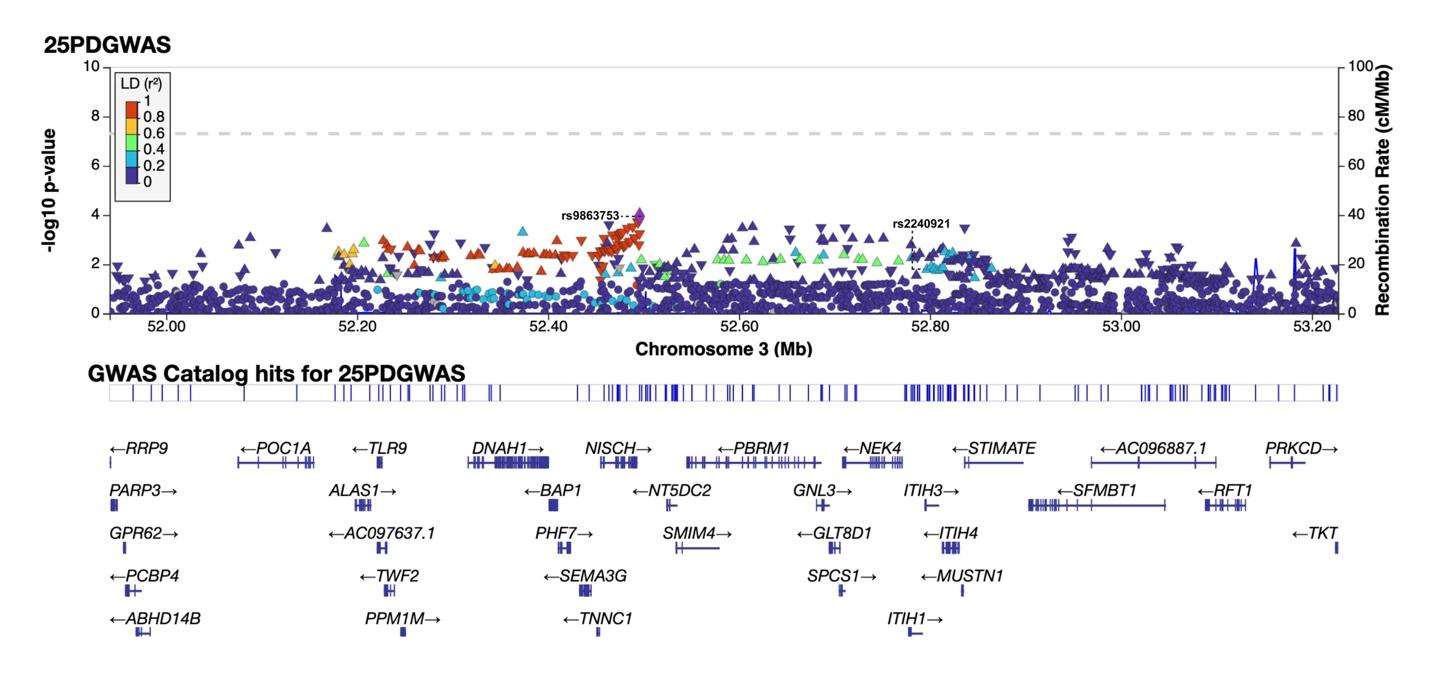

Figure S3b. rs2240921(3:52796748) in 2025 PD GWAS. The Genome build is GRCh38.

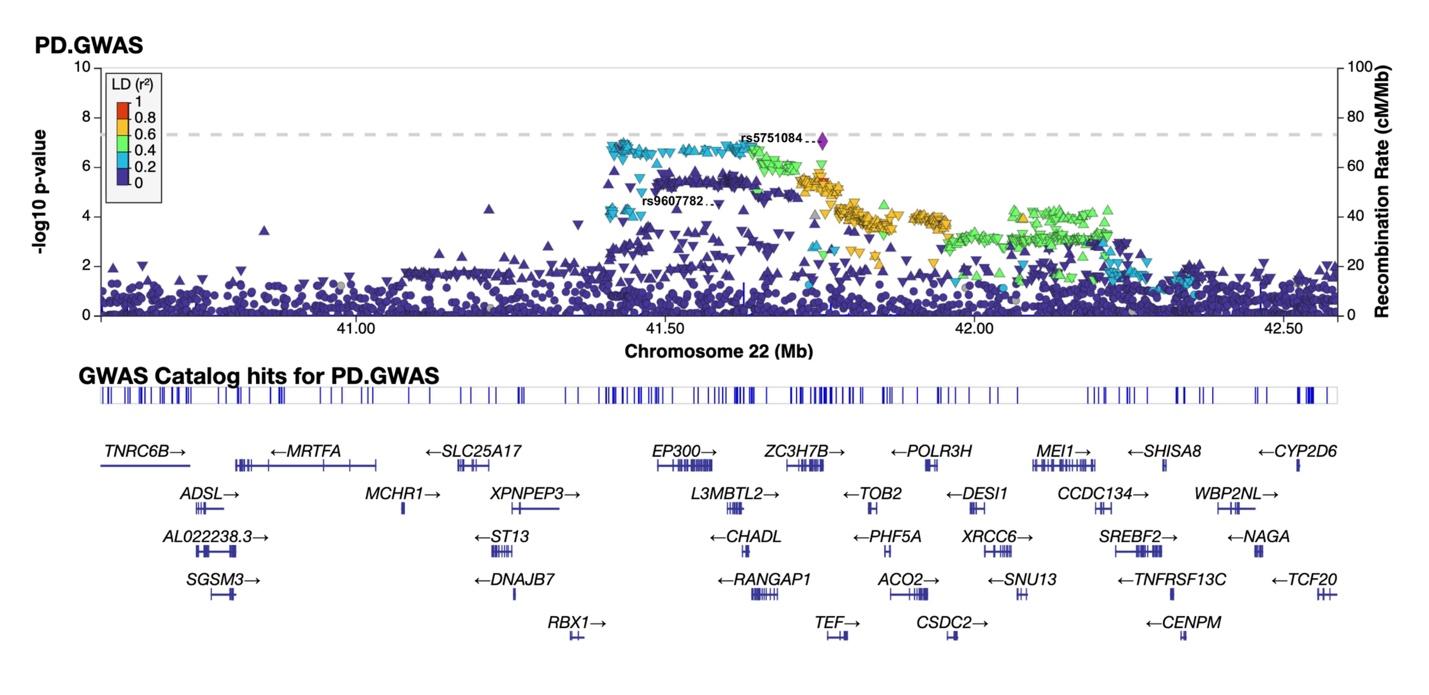
Figure S4a. rs9607782(22:41587556) in 2019 PD GWAS. The Genome build is GRCh37.

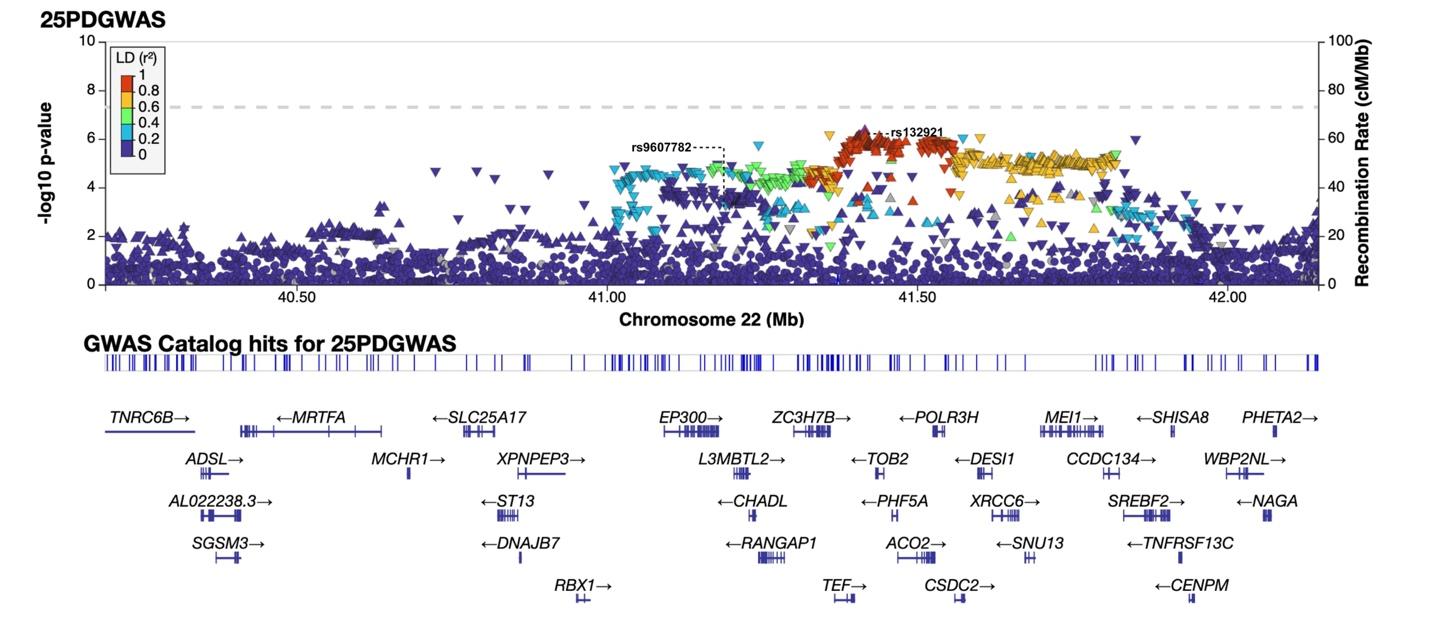

Figure S4b. rs9607782(22:41191552) in 2025 PD GWAS. The Genome build is GRCh38.

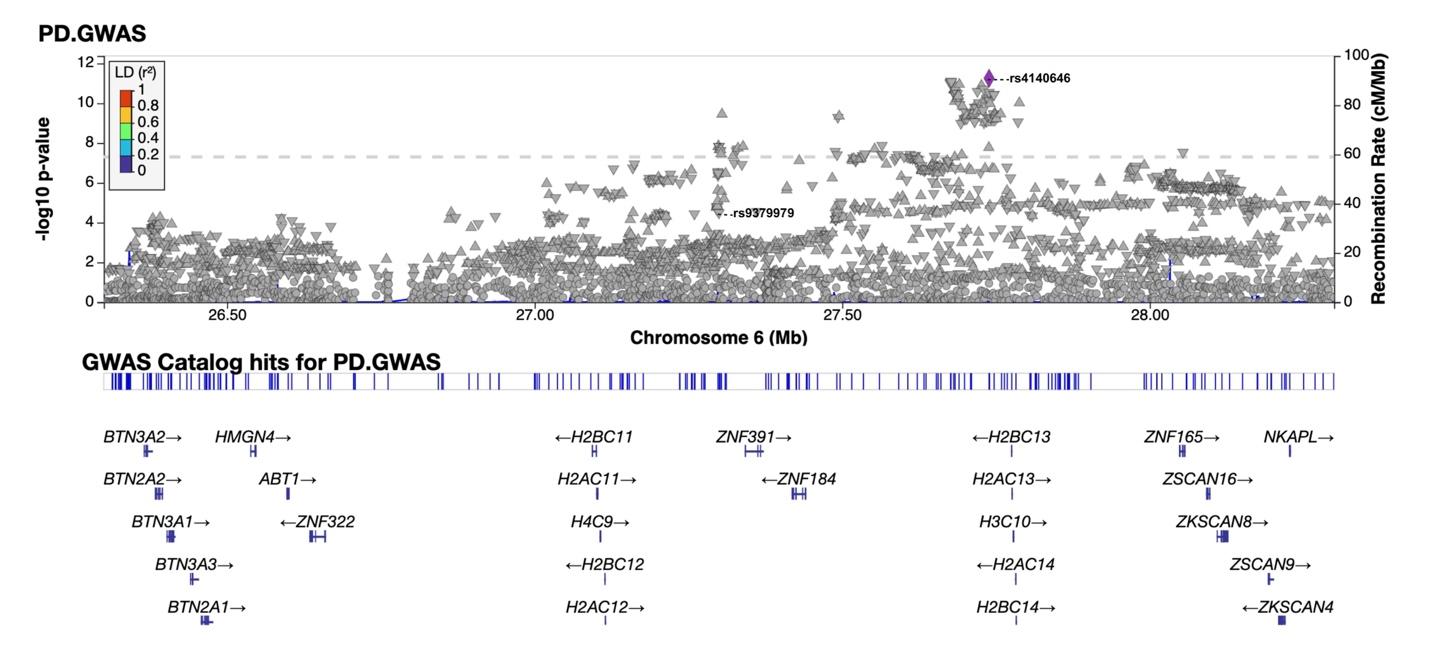

Figure S5a. rs9379979(6:27299614) in 2019 PD GWAS. The Genome build is GRCh37.

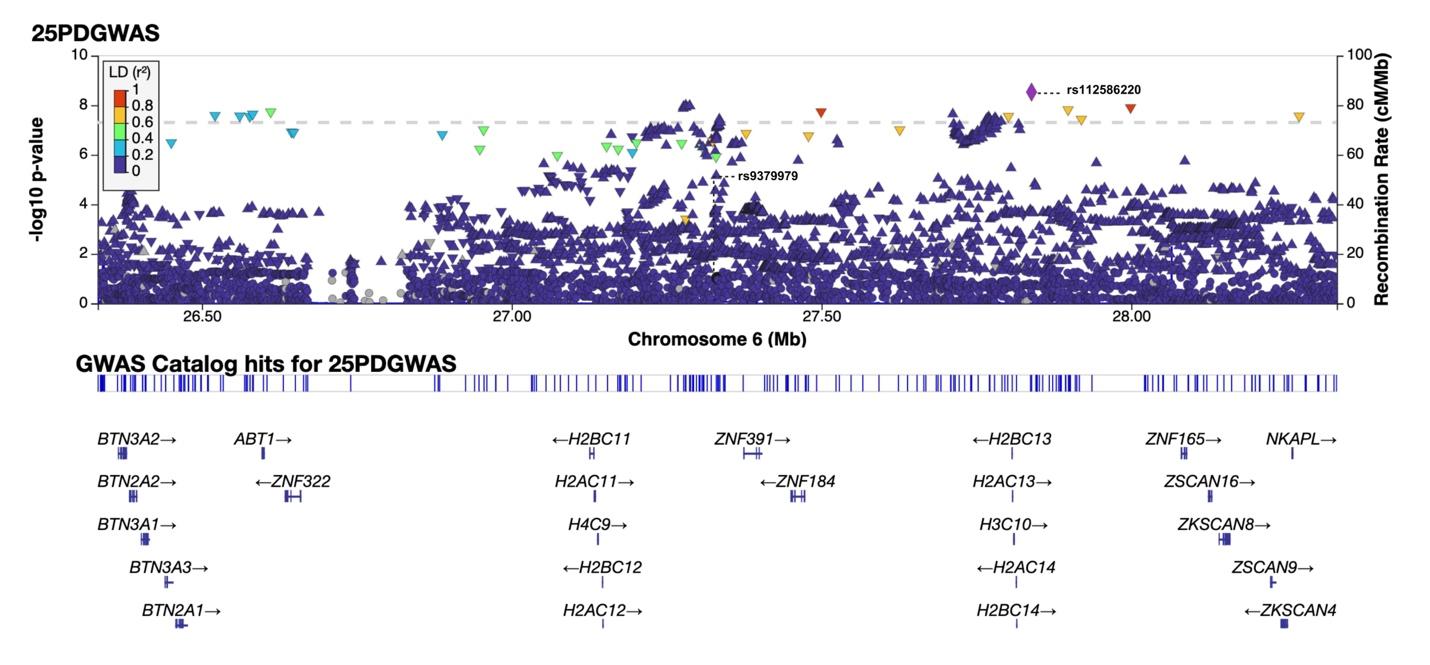

Figure S5b. rs9379979(6:27331835) in 2025 PD GWAS. The Genome build is GRCh38.

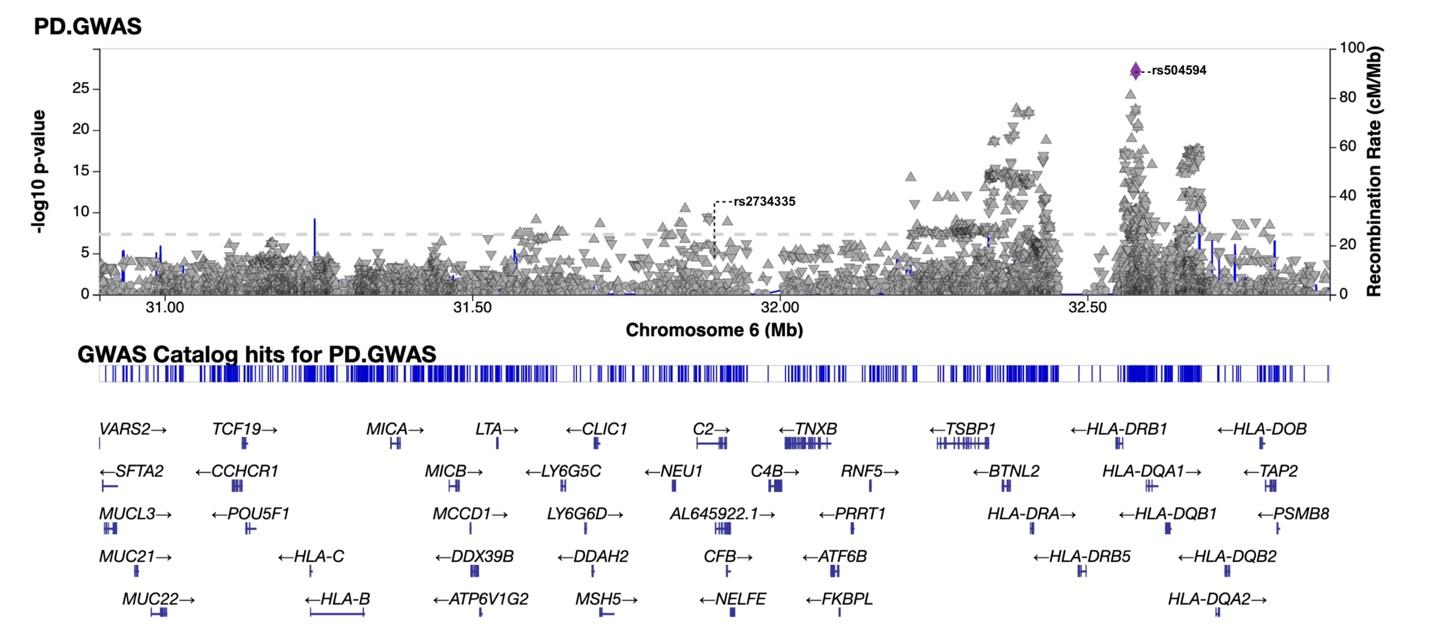

Figure S6a. rs2734335(6:31893944) in 2019 PD GWAS. The Genome build is GRCh37.

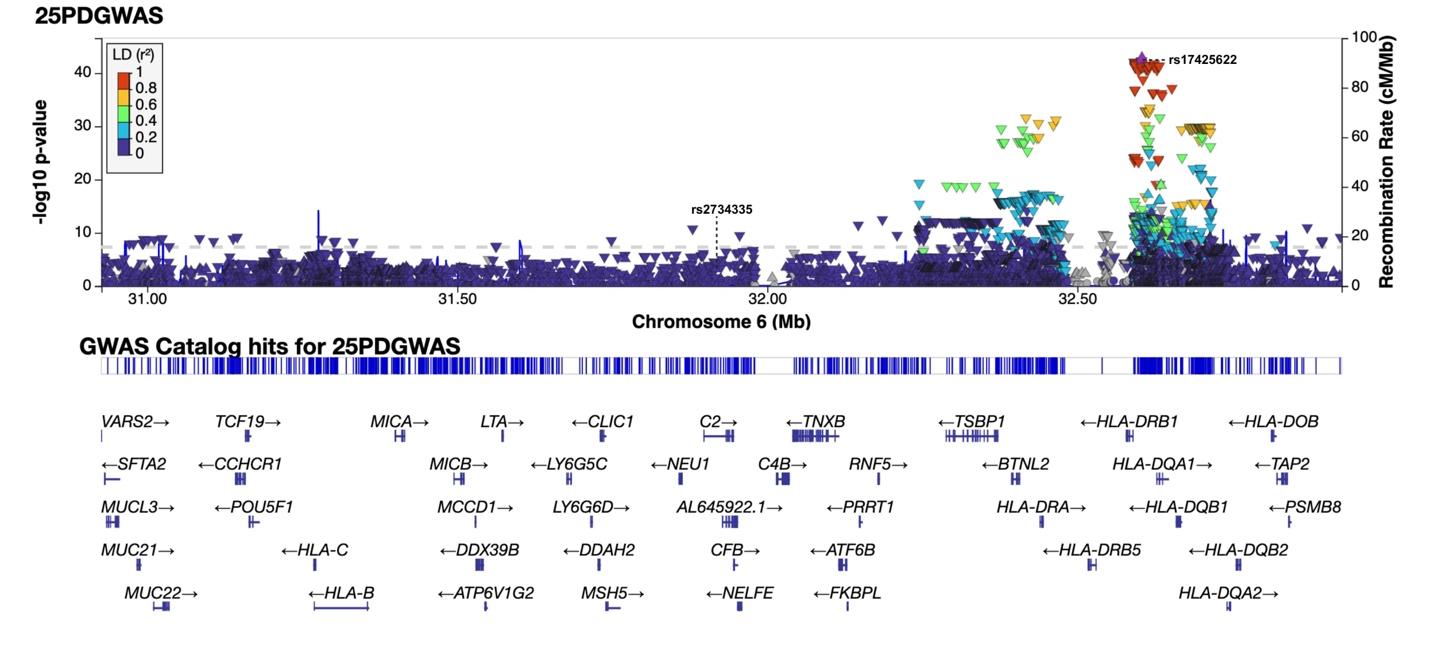

Figure S6b. rs2734335(6:31926167) in 2025 PD GWAS. The Genome build is GRCh38.

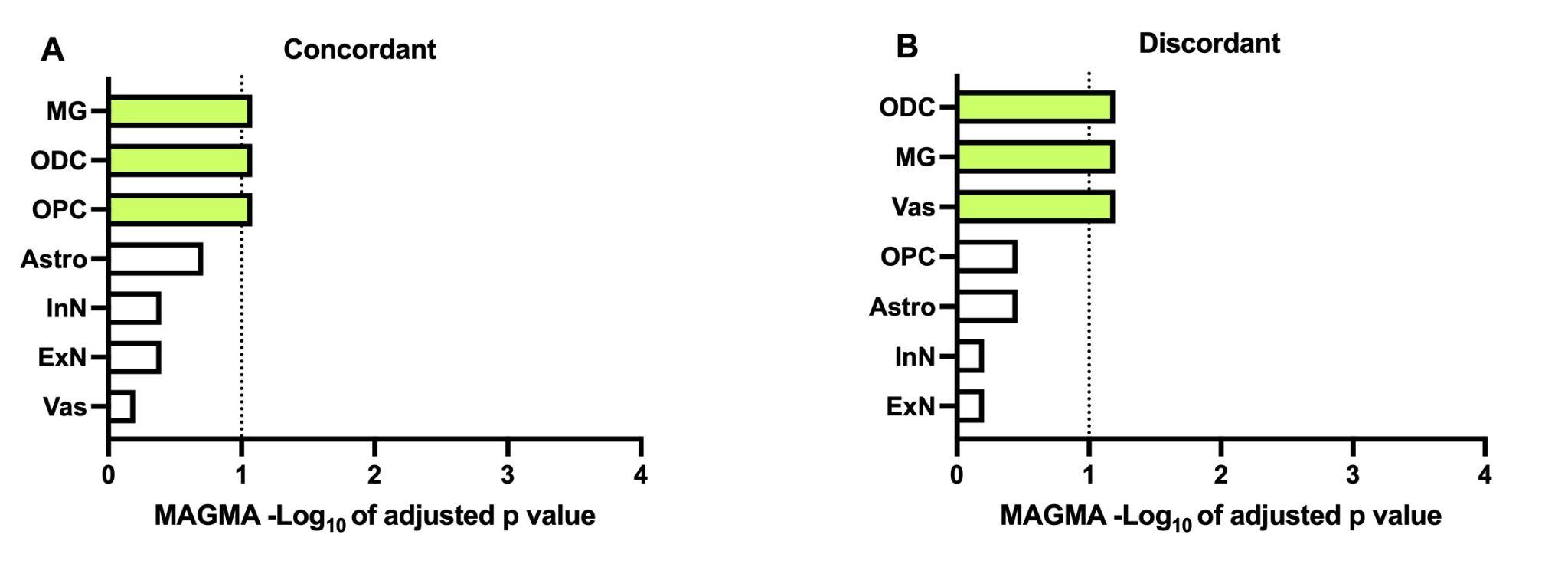

Figure S7. Sanity check was performed after exclusion of pathogenic genes (*SNCA*, *GBA1*, and *LRRK2*) in PD.(A) Multi-marker analysis of genomic annotation (MAGMA) gene set enrichment based on PDconcordant SNP list in postmortem brain issue derived single cell RNA sequencing dataset (PMID: [39138468](https://pubmed.ncbi.nlm.nih.gov/39138468/)). (B) MAGMA gene set enrichment based on PDdiscordant SNP list n postmortem brain issue derived single cell RNA sequencing dataset (PMID: [39138468](https://pubmed.ncbi.nlm.nih.gov/39138468/)). The dotted line indicates the FDR-adjusted significance threshold of 0.1.

Table S1 Top shared SNPs between PD and SZ with a Bayesian statistical approach.

| SNP | ZParkinson | ZSchizo | TotalZ |
| --- | --- | --- | --- |
| rs9379968 | -5.063830 | -6.767775 | 11.831605 |
| rs9270836 | 5.310606 | -5.644901 | 10.955507 |
| rs298602 | -5.623656 | -5.057212 | 10.680868 |
| rs546381 | 5.042553 | -5.104889 | 10.147442 |
| **rs11649804** | 4.902913 | 4.968494 | 9.871406 |

Table S6. Summary of rs11649804 and rs12941356 variants between Neurodegenerative and Psychiatric Disorders.

|  |  | |  | | Neurodegenerative disorders | | | | | | | | |
| --- | --- | --- | --- | --- | --- | --- | --- | --- | --- | --- | --- | --- | --- |
| Disease |  | rs11649804  17:17696755:C:A | | | | | |  |  | rs12941356  17:17716531:A:G | | | |
|  | EA | beta | | SE | | p value | OR (95%CI) |  | EA | beta | SE | p value | OR (95%CI) |
| PD | A | -0.051 | | 0.010 | | 8.49e-07 | 0.95(0.93-0.97) |  | A | -0.051 | 0.0096 | 1.31e-07 | 0.95(0.93-0.97) |
| AD | A | -0.023 | | 0.0088 | | 0.0098 | 0.98(0.96-0.99) |  | A | -0.013 | 0.0082 | 0.11 | 0.99(0.97-1.00) |
| FTD | A | 0.057 | | 0.058 | | 0.33 | 1.06(0.94-1.19) |  | A | -0.00088 | 0.054 | 0.99 | 1.00(0.90-1.11) |
| ALS | A | -0.020 | | 0.013 | | 0.12 | 0.98(0.96-1.01) |  | A | -0.025 | 0.012 | 0.035 | 0.98(0.95-1.00) |
| MSA | A | -0.037 | | 0.059 | | 0.53 | 0.96(0.86-1.08) |  | A | 0.024 | 0.055 | 0.66 | 1.02(0.92-1.14) |
| PSP | A | NA | | NA | | 0.56 | NA |  | A | NA | NA | 0.87 | NA |
|  |  | |  | | Psychiatric Disorders | | | | | | | | |
| Disease |  | rs11649804 | | | | | |  |  | rs12941356 | | | |
|  | EA | beta | | SE | | p value | OR (95%CI) |  | EA | beta | SE | p value | OR (95%CI) |
| SZ | C | -0.047 | | 0.0094 | | 6.66e-07 | 0.95(0.94-0.97) |  | A | 0.036 | 0.0088 | 4.00e-05 | 1.04(1.02-1.05) |
| ADHD | A | 0.029 | | 0.011 | | 0.0068 | 1.03(1.01-1.05) |  | A | -0.027 | 0.0098 | 0.0065 | 0.97(0.95-0.99) |
| AN | C | 0.024 | | 0.015 | | 0.10 | 1.02(0.99-1.05) |  | A | -0.018 | 0.014 | 0.20 | 0.98(0.96-1.01) |
| ASD | A | -0.023 | | 0.016 | | 0.14 | 0.98(0.95-1.01) |  | A | -0.026 | 0.014 | 0.073 | 0.97(0.95-1.00) |
| BP | C | 0.0071 | | 0.015 | | 0.63 | 1.01(0.98-1.04) |  | A | 0.012 | 0.014 | 0.39 | 1.01(0.98-1.04) |
| MDD | A | 0.0056 | | 0.013 | | 0.66 | 1.01(0.98-1.03) |  | A | -0.0073 | 0.012 | 0.54 | 0.99(0.97-1.02) |
| OCD | A | 0.086 | | 0.037 | | 0.018 | 1.09(1.01-1.17) |  | A | 0.067 | 0.034 | 0.050 | 1.07(1.00-1.14) |
| PPD | A | 0.17 | | -0.024 | | 0.017 | 1.19(1.13-1.24) |  | A | 0.29 | -0.015 | 0.014 | 1.34(1.30-1.38) |
| PTSD | A | NA | | NA | | 0.61 | NA |  | A | NA | NA | 0.66 | NA |
| smoking initiation | A | -0.0027 | | 0.0016 | | 0.096 | 0.997 (0.994-1.00) |  | NA | | | | |

Abbreviations: PD, Parkinson’s disease; AD, Alzheimer’s disease; FTD, Frontotemporal dementia; ALS, Amyotrophic lateral sclerosis; MSA, Multiple system atrophy; PSP, Progressive supranuclear palsy; SZ, Schizophrenia; ADHD, Attention-deficit/hyperactivity disorder; AN, Anorexia nervosa; ASD, Autism spectrum disorder; BP, Bipolar disorder; MDD, Major depressive disorder; OCD, Obsessive-compulsive disorder; PPD, Post-Partum Depression; PTSD, Post-traumatic stress disorder; EA, effect allele; OR, odds ratio; CI, confidence interval; SE, standard error.

Table S7. Overview of shared novel genetic variants between PD and SZ identified from the GWAS catalogue.

| SNP | Mapped gene | risk allele | RAF | Beta/OR (CI) | P value | Trait |
| --- | --- | --- | --- | --- | --- | --- |
| rs2240921  (3:52830764:C:T) | *ITIH3* | T | 0.12 | Beta: 0.026 (0.022-0.03) | 1e-20 | high density lipoprotein cholesterol measurement |
|  |  | T | / | Beta: 0.029 (0.020-0.037) | 3e-11 | BMI-adjusted waist-hip ratio |
|  |  | T | / | / | 1e-09 | fatty acid amount |
| rs11649804  (17:17696755:C:A) | *RAI1* | / | / | Beta: 0.13 (0.08-0.18) | 3e-07 | susceptibility to childhood ear infection measurement |
|  |  | A | 0.41 | / | 1e-10 | hematocrit |
|  |  | A | / | Beta: 0.020 (0.012-0.026) | 3e-08 | BMI-adjusted waist-hip ratio |
|  |  | A | 0.29 | Beta: 0.020 (0.015-0.025) | 3e-16 | monocyte count |
| rs9607782  (22:41587556:T:A) | *EP300-AS1* | A | 0.23 | OR: 1.08 (1.06-1.13) | 2e-11 | Schizophrenia |
|  |  | A | / | OR: 1.07 (1.04-1.09) | 9e-09 | autism spectrum disorder, schizophrenia |
|  |  | / | / | Beta: 0.015 (0.011-0.02) | 9e-11 | schizophrenia, intelligence, self-reported educational attainment |
|  |  | / | / | OR: 1.16 (1.12-1.2) | 3e-18 | Alzheimer disease, polygenic risk score |

RAF: allele frequency of risk allele; OR, odds ratio.
