## Supplementary material for "Unraveling the Genetic Overlap Between Parkinson’s Disease and Schizophrenia Through Genome-wide Association and Cell-Type Specific Transcriptomic Analysis": Cohort overview of PD and SZ GWAS

##### Cohorts’ information of PD GWAS (Nalls et al., 2019) [1]

| Study | Ncase | Ncontrol |
| --- | --- | --- |
| 1. 23andMe, post-Chang et al. 2017 enrollment | 2,448 | 571,411 |
| 2. Baylor College of Medicine / University of Maryland | 769 | 195 |
| 3. Finnish Parkinson's | 386 | 493 |
| 4. Harvard Biomarker Study (HBS) | 527 | 472 |
| 5. McGill Parkinson's | 582 | 905 |
| 6. Oslo Parkinson's Disease Study | 476 | 462 |
| 7. Parkinson's Disease Biomarker's Program (PDBP) | 512 | 282 |
| 8. Parkinson's Progression Markers Initiative (PPMI) | 363 | 165 |
| 9. System Genomics of Parkinson's Disease (SGPD) | 1,169 | 968 |
| 10. Spanish Parkinson's (IPDGC) | 2,110 | 1,333 |
| 11. Tubingen Parkinson's Disease cohort (CouragePD) | 666 | 542 |
| 12. Vance (dbGap phs000394) | 620 | 299 |
| 13. UK PDMED (CouragePD) | 1,025 | 655 |
| 14. UK BioBank | 18,618 | 436,419 |
| 15. IPDGC (Nalls et al. 2014 discovery phase)/PDGene | 13,708 | 95,282 |
| 16. NeuroX - dbGaP (phs000918.v1.p1) from IPDGC | 5,851 | 5,866 |
| 17. Parkinson's Disease Web-based Study (PDWBS)/23andMe | 6,476 | 302,042 |
| PD GWAS(Nalls et al., 2019) | 56,306 | 1,417,791 |

Cohorts’ information of SZ GWAS (Trubetskoy V, et al., 2022) [2]

| Dataset | Platform | Study | Ncase | Ncontrol |
| --- | --- | --- | --- | --- |
| scz_xume2_eur_sr-qc | OMEX | •Adolfsson, R \| Umeå, Sweden  •Betula study, PGC | 595 | 1,638 |
| scz_xtop8_eur_sr-qc | A6.0 | •Andreassen, O \| Norway (TOP)  •Thematically Organised Psychosis Research (TOP) study | 377 | 403 |
| scz_to10c_eur_sr-qc | OMEX | •Andreassen, O \| Norway  •TOP study and local hospital[3] | 970 | 5,040 |
| scz_eusp2_eur_sr-qc | COEX | Arango, C \| Spain(EUGEI)[4-5] | 338 | 490 |
| scz_celso_eur_sr-qc | PSYC | •Arango, C \| Spain  •CIBERSAM (Network Biomedical Research Centre in Mental Health); PGC | 2,030 | 1,517 |
| scz_eutu2_eur_sr-qc | COEX | •Atbaşoğlu, EC; Saka, M \| Turkey  •EUGEI [6-7] (http://www.eu-gei.eu) | 393 | 690 |
| scz_geba1_eur_sr-qc | PSYC | Baune, B \| Germany | 397 | 703 |
| scz_sb2aa_eur_sr-qc | OMEX | •Belangero, S \| Brazil  •Schizophrenia and First Episode of Psychosis Program at UNIFESP, Sao Paulo, Brazil[8-9] | 250 | 237 |
| scz_xedin_eur_sr-qc | A6.0 | •McIntosh, A \| Edinburgh, UK  •International Schizophrenia Consortium studies[10] | 368 | 284 |
| scz_xaarh_eur_sr-qc | I650 | Børglum, A \| Denmark | 883 | 873 |
| scz_cogs1_eur_sr-qc | PSYC | •Braff, D \| USA  •Consortium on the Genetics of Schizophrenia (COGS)[11] | 428 | 476 |
| scz_xpewb_eur_sr-qc  (excluded) | I1M | Bramon \| Spain (PEIC, WTCCC2) https://www.wtccc.org.uk/ccc2/wtccc2_studies.html | 641 | 1,892 |
| scz_xpews_eur_sr-qc  (excluded) | I1M | Bramon \| Seven countries (PEIC, WTCCC2) | 150 | 236 |
| scz_xmsaf_eur_sr-qc | A6.0 | Buxbaum, J \| New York, US & Israel | 327 | 139 |
| scz_rouin_eur_sr-qc | PSYC | Campion, D; Laurent-Levinson, C \| France | 204 | 185 |
| scz_xdubl_eur_sr-qc | A6.0 | Corvin, A \| Ireland [10] | 272 | 860 |
| scz_du2aa_eur_sr-qc | COEX | •Corvin, A; Morris, D \| Ireland  •PGC, EUEGI, Ireland, CardiffCOGS study[12] | 345 | 245 |
| scz_xirwt_eur_sr-qc  (excluded) | A6.0 | Corvin, A; Riley, B \| Ireland (WTCCC2) [13], WTCCC2 as replication sample | 1,309 | 1,022 |
| scz_gap1a_eur_sr-qc | COEX | •Di Forti, M \| London, UK  •Genetic and Psychotic Disorders Study case-control project[14] | 152 | 164 |
| scz_enric_eur_sr-qc | PSYC | Domenici, E \| Italy | 700 | 574 |
| scz_xgras_eur_sr-qc | AXI | Ehrenreich, H \| Germany (GRAS) | 1,086 | 1,232 |
| scz_xegcu_eur_sr-qc | omni | •Esko, T \| Estonia (EGCUT)  •Estonian Genome Project of University of Tartu (EGCUT)[15] | 239 | 1,177 |
| scz_xjrsa_eur_sr-qc | I1M | •Esko, T; Li, Q; Malhotra D \| J&J and Roche cases, EGCUT controls; •Genotype data: Estonian Biobank; •https://genomics.ut.ee/en/access-biobank | 1,181 | 2,313 |
| scz_xjr3a_eur_sr-qc | I317 |  | 362 | 325 |
| scz_xjr3b_eur_sr-qc | I317 |  | 647 | 639 |
| scz_xjri6_eur_sr-qc | I610 |  | 260 | 130 |
| scz_price_eur_sr-qc | PSYC | •Gareeva, A; Khusnutdinova, E \| Ufa, Russia  •GWAS analysis in Russian SZ sample from the Volga Ural region | 841 | 727 |
| scz_gawli_eur_sr-qc | PSYC | •Gawlik, M \| Germany  •Genetics of Psychoses | 1,255 | 1,555 |
| scz_xmgs2_eur_sr-qc | A6.0 | •Gejman, P \| US, Australia (MGS)  •Molecular Genetics of Schizophrenia (MGS) collaboration[16]  •A survey company (Knowledge Networks, under MGS guidance) collected controls[17] | 2,681 | 2,653 |
| scz_mosc2_eur_sr-qc | I650 | Golimbet, V \| Moscow[18] | 410 | 433 |
| scz_xuclo_eur_sr-qc | A6.0 | •McQuillin, A \| London, UK[10]  •Schizophrenia 20.21 resource at the NIMH Repository & Genomics Resource: https://www.nimhgenetics.org/download-tool/SZ | 521 | 494 |
| scz_xersw_eur_sr-qc | omni | •Jönsson, E \| Sweden (Hubin) [19]  •Case(SGENE-plus)  •Control(Karolinska Institute, Stockholm County) | 322 | 332 |
| scz_xbuls_eur_sr-qc | A6.0 | •Kirov, G \| Bulgaria  •control[10] | 195 | 608 |
| ms.scz_xuktr_eur_sr-qc | omni | Kirov, G; Owen, M \| Bulgaria | 70 | 140 |
| ms.scz_xbutr_eur_sr-qc | A6.0 | Kirov, G; Owen M \| Bulgaria | 741 | 1,156 |
| scz_xlktu_eur_sr-qc | A6.0 | •Kennedy JL, Collier DA \| Canada, US(Lilly), US (MIGen)  •Myocardial Infarction Genetics Consortium (MIGen, dbGaP ID phs000294.v1.p1) [20] | 322 | 332 |
| scz_paris_eur_sr-qc | PSYC | Krebs, M \| France, PSYDEV collection(case and control) | 316 | 390 |
| scz_xajsz_eur_sr-qc | omni | •Lencz, T; Darvasi A \| Israel[21]  •Ashkenazi Jewish repository (Hebrew University Genetic Resource, http://hugr.huji.ac.il) | 896 | 1,595 |
| scz_xlacw_eur_sr-qc  (excluded) | I550 | •Levinson, D \| 22885689 \| Six countries, WTCCC controls[23]  •a larger pedigree-based study [22] | 157 | 466 |
| ms.scz_xlemu_eur_sr-qc | I650 | Levinson, D \| Six countries[22] | 585 | 0 |
| scz_xzhh1_eur_sr-qc | A500 | Malhotra, A \| New York, US[24] | 191 | 190 |
| scz_mcqul_eur_sr-qc | PSYC | •McQuillin, A \| United Kingdom[25]  •DNA Polymorphisms in Mental Illness (DPIM) study  • 480 ECACC Human Random Control (HRC) samples obtained from Public Health England(supplemented control) | 1,351 | 1,310 |
| scz_xasrb_eur_sr-qc | I650 | •Mowry, B; Morgan, V \| Australia  •Australian Schizophrenia Research Bank  •Survey of High Impact Psychosis (SHIP) study[26] | 509 | 310 |
| scz_viyo1_eur_sr-qc | PSYC | Nimgaonkar, V \| USA[27] | 356 | 131 |
| scz_braz2_eur_sr-qc | COEX | •Menezes, P; Belangero, S \| Brazil [5]  •EUGEI | 110 | 334 |
| scz_xcaws_eur_sr-qc  (excluded) | A500 | •O'Donovan, M; Owen, M \| Cardiff, UK[28]  •WTCCC(control) [23] 1958 British Birth Cohort and a panel of consenting blood donors (UK Blood Service) | 424 | 306 |
| scz_xclm2_eur_sr-qc | I1M | •Walters, J; O’Donovan, M; Owen M \| UK (CLOZUK)  •CLOZUK(case) | 3,466 | 4,297 |
| scz_xclo3_eur_sr-qc  (excluded) | omni | •WTCCC2(control), UK National Blood Transfusion Service | 2,150 | 2,083 |
| scz_xucla_eur_sr-qc | I550 | Ophoff, R \| Netherlands[19] | 705 | 637 |
| scz_xfi3m_eur_sr-qc | I317 | •Palotie, A \| Finland  •THL Psychiatric Family Collections | 186 | 930 |
| scz_xfii6_eur_sr-qc | I550 | •the Finnish Health 2000 survey, the National FINRISK Study(control), Northern Finland Birth Cohort (NFBC)(control), Helsinki Birth Cohort Study (HBCS)(control) | 361 | 1,082 |
| scz_xport_eur_sr-qc | A6.0 | Pato, C \| Portugal[10] | 346 | 216 |
| scz_gpc2a_eur_sr-qc | PSYC | •Pato, C \| Multiple sites  •Genomic Psychiatry Cohort (GPC)[29] | 1,957 | 2,062 |
| scz_xcims_eur_sr-qc | ill | Petryshen, T \| Boston, US (CIDAR) | 71 | 69 |
| scz_xboco_eur_sr-qc | I550 | •Rietschel, M; Rujescu, D; Nöthen, M \| Bonn/Mannheim, Germany;  •MooDS Consortium in Mannheim[19], Bonn[19], Munich and Jena  •three population-based epidemiological studies (PopGen)(control) [30], the Cooperative Health Research in the Region of Augsburg (KORA) study[31], the Heinz Nixdorf Recall (HNR) study[32] | 1,847 | 2,170 |
| scz_bep1b | GSA | •Ripke, S \| Berlin  •Berlin Psychosis Study (BePS) | 294 | 573 |
| scz_sanch_eur_sr-qc | PSYC | •Cervilla, M \| Granada  •GENIMS[33] (case)  •PISMA[34] (control) | 335 | 1,209 |
| scz_xmunc_eur_sr-qc | I317 | •Rujescu, D \| Munich, Germany[19] | 437 | 351 |
| scz_serri_eur_sr-qc | PSYC | •Serretti, A \| Italy | 217 | 238 |
| scz_xaber_eur_sr-qc | A6.0 | St Clair, D \| Aberdeen, UK[10] | 720 | 699 |
| scz_xcati_eur_sr-qc | A500 | •Sullivan, PF \| US (CATIE)  •Clinical Antipsychotics Trials of Intervention Effectiveness (CATIE) project[35-37]  •MGS  •Schizophrenia 20.21 resource, study 17, at the NIMH Repository & Genomics Resource: https://www.nimhgenetics.org/download-tool/SZ | 409 | 392 |
| scz_xswe1_eur_sr-qc | A5.0 | •Sullivan, PF; Sklar P; Hultman C \| Sweden; •Swedish Schizophrenia Study[38] , sw1-2[10], sw1-4[39] ; •Schizophrenia 20.21 resource at the NIMH Repository & Genomics Resource: https://www.nimhgenetics.org/download tool/SZ. | 221 | 214 |
| scz_xs234_eur_sr-qc | A6.0 |  | 2,077 | 2,341 |
| scz_xswe5_eur_sr-qc | omni |  | 1,801 | 2,617 |
| scz_xswe6_eur_sr-qc | omni |  | 1,094 | 1,219 |
| ms.scz_grtr1_eur_sr-qc | COEX | •Van Os, J \| Netherlands and Belgium | 145 | 290 |
| scz_gro2a_eur_sr-qc | COEX | •GROUP STUDY(case) [40] | 329 | 277 |
| scz_eu5me_eur_sa-qc | COEX | Van Os, J; O’Donovan, M \| EUGEI [4-5] | 615 | 182 |
| scz_cgs1c_eur_sr-qc | OMEX | •Walters, J \| UK  •CardiffCOGS2 study | 524 | 3,115 |
| scz_xcou3_eur_sr-qc | omni | •Walters, J \| Cardiff, UK  •CardiffCOGS1 study[41] | 540 | 693 |
| scz_xdenm_eur_sr-qc | I650 | •Werge, T \| Denmark | 492 | 458 |
| scz_clz2a_eur_sr-qc  (excluded) | OMEX | •Walters, J; O’Donovan, M; Owen, M \| CLOZUK[42]  •4641 controls in WTCCC2 | 5,370 | 6,940 |
| scz_xlie5_eur_sr-qc | I550 | •Weinberger, D \| NIMH CBDB  •Clinical Brain Disorders Branch of the NIMH ‘Sibling[43]  •Schizophrenia 20.21 resource at the NIMH Repository & Genomics Resource | 509 | 389 |
| scz_xlie2_eur_sr-qc | O25 | •https://www.nimhgenetics.org/download-tool/SZ | 137 | 269 |
| scz_xpfla_eur_sr-qc | I550 | •Paciga, S \| Pfizer \| Multiple countries  •cases from seven multi-centre randomized, double-blind efficacy and safety clinical trials (A1281063, A1281134, A1281148, A245-102, NRA7500001, NRA7500002, NRA7500003, and NRA7500004) as well as a set of purchased samples (NRA9000099)  •The controls (A9011027) were recruited in a multi-site, cross-sectional, non-treatment prospective trial in US  •LEADe[44] and UCSD MCI[45] | 681 | 1,174 |
| SZ GWAS (Trubetskoy V, et al., 2022) | | | 55,193 | 74,132 |
